# ALPaCA: Adapting Llama for Pathology Context Analysis to enable slide-level question answering

**DOI:** 10.1101/2025.04.22.25326190

**Authors:** Zeyu Gao, Kai He, Weiheng Su, Ines P. Machado, William McGough, Mercedes Jimenez-Linan, Brian Rous, Chunbao Wang, Chengzu Li, Xiaobo Pang, Tieliang Gong, Ming Y. Lu, Faisal Mahmood, Mengling Feng, Chen Li, Mireia Crispin-Ortuzar

## Abstract

Large Vision Language Models (LVLMs) have recently revolutionized computational pathology. LVLMs transform pathology image embeddings into tokens recognizable by large language models, facilitating zero-shot image classification, description generation, question answering, and interactive diagnostics. In clinical practice, pathological assessments often require the analysis of entire tissue slides, integrating information from multiple sub-regions and magnification levels. However, existing LVLM frameworks have been restricted to the analysis of small, predefined regions of interest, lacking the ability to analyze pyramidal, gigapixel-scale whole-slide images (WSIs). In this work, we introduce **ALPaCA** (**A**dapting **L**lama for **Pa**thology **C**ontext **A**nalysis), and train the first general-purpose slide-level LVLM, leveraging 35,913 WSIs with curated descriptions alongside 341,051 question and answer pairs encompassing diverse diagnoses, procedures, and tissue types. By developing LongFormer, a vision-text interactive slide-level adaptor, and integrating it with a Gaussian mixture model-based prototyping adaptor, followed by training with Llama3.1, ALPaCA achieves superior performance in slide-level question answering, achieving over 90% accuracy in close-ended tests and high accuracy in open-ended questions as evaluated by expert pathologists, highlighting its potential for slide-level computer-aided diagnosis systems. Additionally, we show that ALPaCA can be readily fine-tuned on in-depth, organ-specific, or disease-specific datasets, underscoring its adaptability and utility for specialized pathology tasks.

## 1 Introduction

In recent years, computational pathology has evolved rapidly, largely due to progress in deep learning and digital scanning technology [1]. These breakthroughs have made it possible to automatically analyse whole-slide images (WSIs), supporting tasks like tumor morphological classification [2–5], biomarker prediction [6–8], expression prediction [9–11], treatment response [12–14] and prognostic analysis [15–17]. Notably, the rise of foundational models for pathology—trained on extensive collections of unlabeled WSIs—has introduced task-agnostic model backbones capable of learning broad and generalizable representations [18–22]. This innovation significantly enhances performance and label efficiency across diverse computational pathology tasks. However, most of these models disregard textual pathology knowledge and still rely on predefined tasks that require trainable heads for downstream applications, limiting their capacity for interactive reasoning and practical diagnostic use.

The rise of large language models (LLMs) [23–25] has reshaped numerous domains of artificial intelligence by achieving unprecedented levels of language understanding and generation. Building on this foundation, Large Vision-Language Models (LVLMs) [26–29] have emerged as multimodal extensions of LLMs, integrating visual information with textual reasoning to enable tasks such as zero-shot image classification, captioning, question answering, and interactive reasoning. Previous works [30–32] have demonstrated the potential of LVLMs in medical imaging diagnosis and research. Among them, pathology-focused LVLMs [33–36] are opening up a new frontier in computational pathology, enabling pathology descriptions, interactive diagnostics, and even follow-up immunohistochemistry (IHC) recommendations for H&E-stained pathology images. Fundamentally, these LVLMs follow a similar framework: pathology image embeddings, extracted using a pre-trained frozen domain-specific encoder, are converted into LLM-recognizable tokens through a simple linear adaptor and concatenated with text tokens for LLM fine-tuning and inference. However, this framework is restricted to region-of-interest (ROI) analysis, focusing on small, predefined areas in isolation, and lacks the capability for comprehensive slide-level analysis. In real-world clinical practice, pathological diagnostics often demand reasoning across entire slides, integrating information from multiple sub-regions and magnification levels. This presents significant challenges for LVLMs that would need adaptors designed to handle the scale, complexity, and hierarchical structure of WSIs efficiently.

To address this gap, we introduce **ALPaCA** (**A**dapting **L**lama for **Pa**thology **C**ontext **A**nalysis), a slide-level LVLM designed for WSI question-answering (QA) across diverse cancer types and tissue sites. While recent efforts have explored slide-level vision-language models, most lack integration with powerful LLMs and focus on generating simple slide descriptions [37–39] or QA for a single cancer type [40]. ALPaCA leverages frozen CONCH [41], a vision encoder pre-trained on 1.17 million pathology image-caption pairs, to extract hierarchical patch embeddings. To enable comprehensive WSI analysis, ALPaCA incorporates various vision-only and vision-text interactive slide-level adaptors to explore the most effective method for compressing patch embeddings into WSI tokens and integrating them with the state-of-the-art publicly available LLM (Llama3.1-8B).

ALPaCA is trained on a general slide caption dataset and a general slide QA dataset sequentially, both curated from the largest publicly available WSI databases, The Cancer Genome Atlas (TCGA) and The Genotype-Tissue Expression (GTEx), with each slide paired to a pathology report or note. For evaluation, 337 hold-out test slides with 4,044 slide-level question and answer (Q&A) pairs spanning different diagnostic types and tissue sites were used, showcasing ALPaCA’s superior performance in general pathology understanding and reasoning. Moreover, fine-tuning on two disease-specific QA datasets separately demonstrates ALPaCA’s adaptability and flexibility for specialized slide-level pathology QA tasks.

## 2 Results

### 2.1 A slide-level large vision language model for pathology question answering

We propose ALPaCA, a comprehensive framework for integrating image-level foundation models with powerful LLMs for pathology slide analysis, while also establishing a systematic approach for slide-level QA evaluation (**Fig. 1 and Section 4**). To develop a slide-level LVLM capable of addressing both general and specific pathology QA tasks, we introduced three key innovations. Firstly, we curated two large-scale datasets, “General Slide-Caption” and “General Slide-QA”, sourced from TCGA and GTEx, comprising 35,913 WSIs with corresponding descriptions and 341,051 Q&A pairs. Secondly, we trained multiple ALPaCA variants with various slide-level adaptors to effectively integrate the pathology foundation model (CONCH) with the Llama3.1 LLM using a sequential training strategy on the curated datasets. In particular, six different slide-level adaptors were developed to explore optimal integration approaches, including four vision-only adaptors—Sampling, Attention-Based Multi-Instance Learning (ABMIL), KMeans, and Gaussian Mixture Model (GMM)—and two vision-text interactive adaptors, QFormer and LongFormer. The best-performing variant, ALPaCA-Hybrid, incorporates LongFormer and GMM, where LongFormer specializes in extracting question-specific detailed local visual representations, while GMM effectively captures global visual information for addressing coarse-grained questions. Finally, we fine-tuned ALPaCA-Hybrid on two disease-specific QA datasets, each tailored to address in-depth morphological and molecular questions related to the corresponding disease type. More details can be found in **Section 4.1-4.4**.

**Fig. 1:**
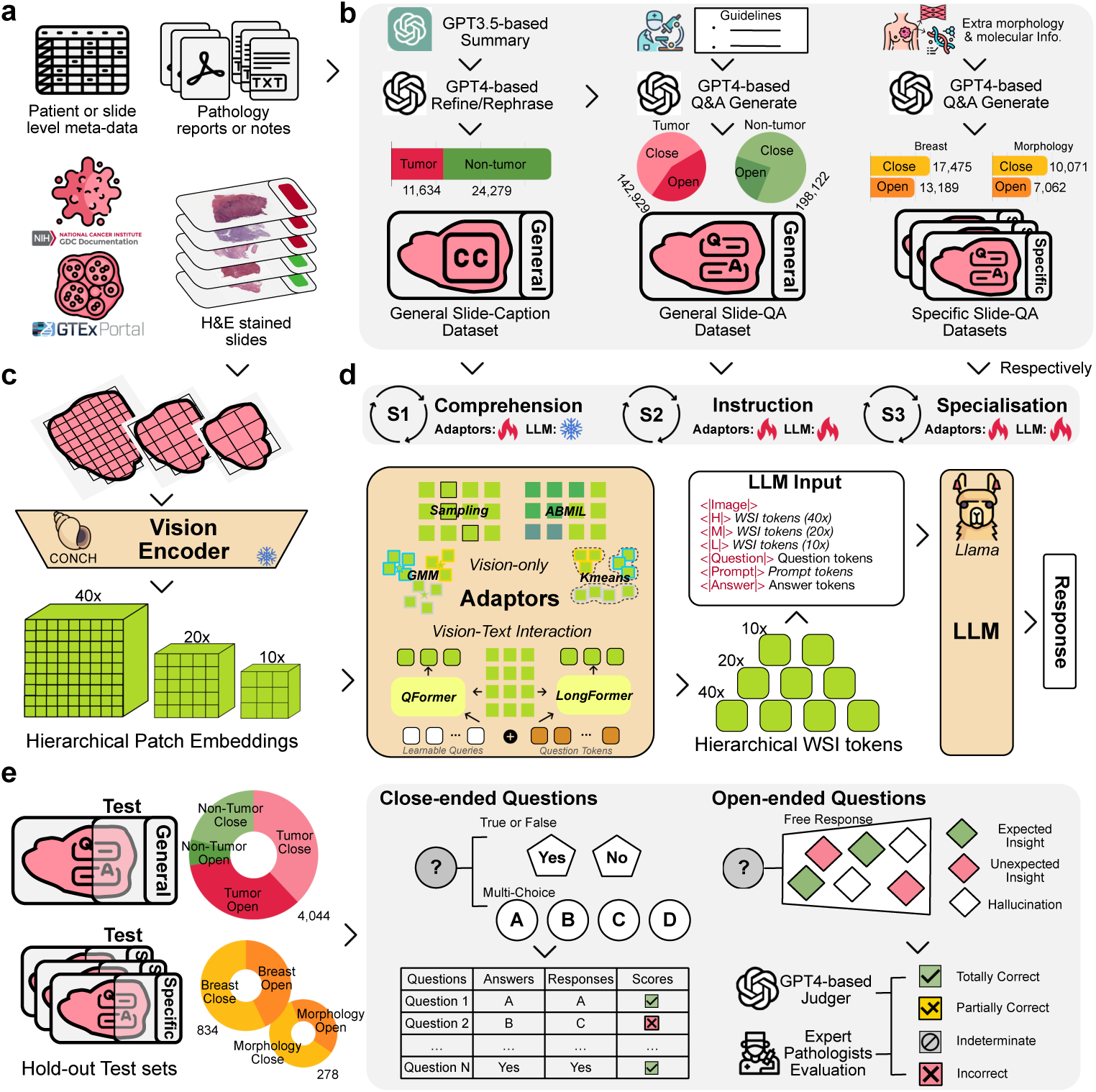
Study overview. **a**, Source data from TCGA (tumor) and GTEx (non-tumor), including metadata, pathology reports/notes, and digitalised H&E-stained slides. **b**, Dataset curation. Pathology reports were summarized with GPT-3.5, and slide descriptions were generated by GPT-4 using summaries/notes, and metadata to form the “General Slide-Caption” dataset. Question-answer pairs were created by GPT-4 with pathologist guidelines for the “General Slide-QA” dataset. Moreover, incorporating extra morphological and molecular information for a subset of slides, the “Specific Slide-QA” datasets were curated. **c**, Hierarchical patch embeddings were extracted by the CONCH encoder at different magnifications. **d**, ALPaCA overview. Patch embeddings were transformed into WSI tokens by various adaptors and combined with text tokens in a structured format as input for the LLM. Training proceeded in three stages—Comprehension, Instruction, and Specialisation—using curated datasets in sequence. **e**, ALPaCA evaluation. Performance was assessed with hold-out test sets for general and specific QA. Close-ended Q&A accuracy was calculated directly, while open-ended Q&A was evaluated with a GPT-4-based judger and expert pathologists.

To assess the performance of ALPaCA variants, we allocated a hold-out test set of 337 slides, covering all diagnostic types and tissue sites in the curated datasets, with separation ensured at the patient level for TCGA and at the case level for GTEx. This test set was strictly excluded from all training stages to prevent data leakage and ensure unbiased evaluation. As a result, evaluation was conducted using 4,044 slide-level Q&A pairs to assess the general pathology QA capability of all ALPaCA variants and 865 and 297 slide-level Q&A pairs, respectively, to evaluate the performance of ALPaCA-Hybrid in two specific pathology QA scenarios. Both automated and expert pathologist assessments were carried out in this study to ensure a comprehensive and unbiased evaluation (**Section 4.5**).

### 2.2 Performance on close-ended questions

We began by evaluating the capabilities of ALPaCA variants on the close-ended general pathology QA task. The test set spanned 32 cancer projects in TCGA and 40 tissue sites in GTEx, with 1,467 questions covering 9 slide examination categories for tumor cases and 936 questions covering 8 histopathological analysis categories for non-tumor cases. In this evaluation, both True/False questions and Multiple-Choice questions allow for a direct comparison between the model’s responses and the ground truth to assess accuracy. For True/False questions, the model is required to correctly follow the instructions and respond with “yes” or “no”. For Multiple-Choice questions, the model must select one answer from the four provided options (A, B, C, or D) and respond accordingly. The accuracy of the model’s responses is calculated based on whether the model’s output matches the correct answer. For the showcase in **Fig. 2a**, ALPaCA-Hybrid correctly answered three out of four questions. However, for Q3, “Is the left fallopian tube involved with the tumor?” the response was incorrect, as the current slide represents ovarian tissue and does not include any fallopian tube structures.

**Fig. 2:**
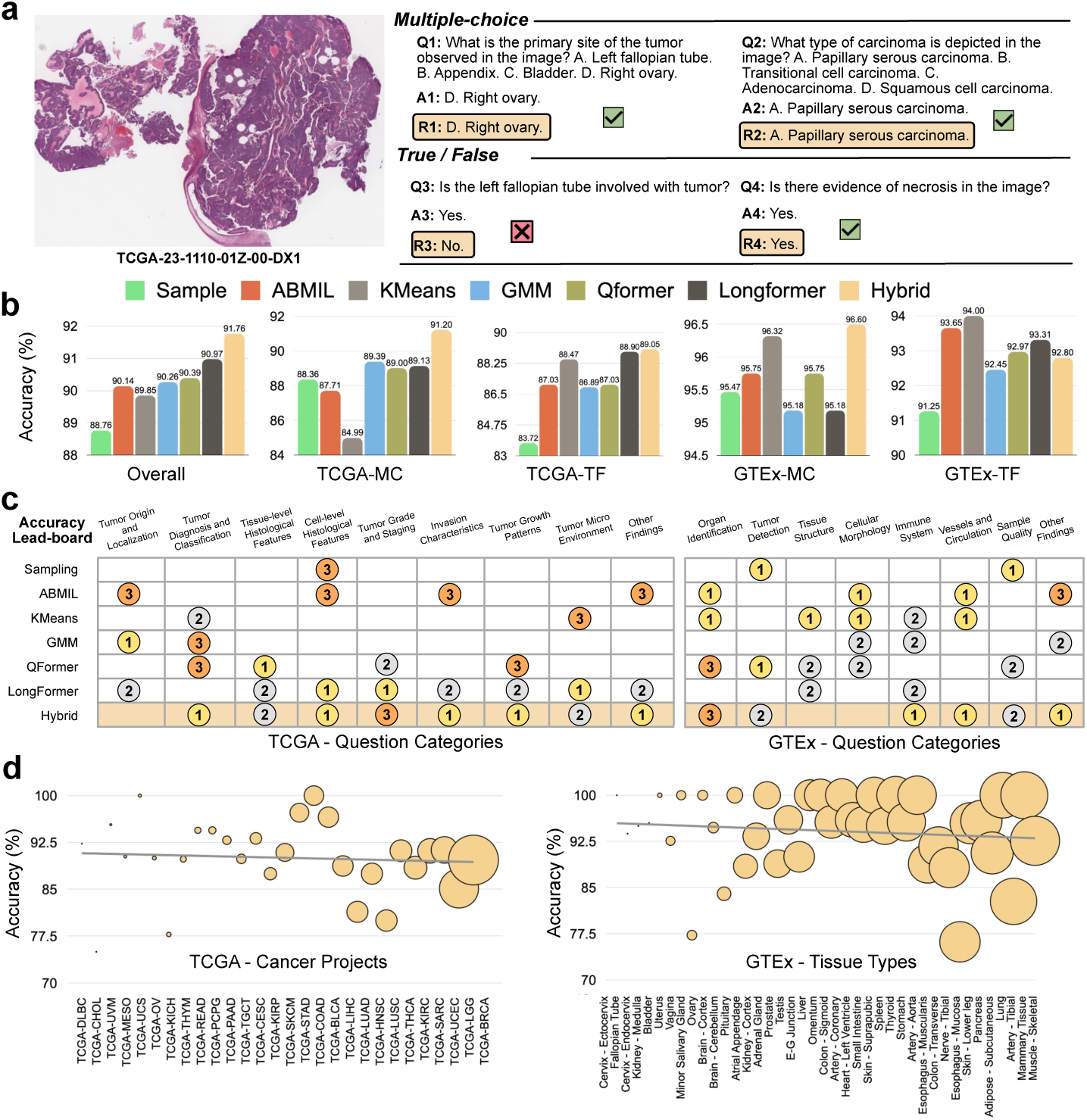
ALPaCA performance on close-ended questions. **a**, An example case of an ovarian tumor slide with corresponding multiple-choice and True/False questions, ground-truth answers, and ALPaCA-Hybrid’s responses. **b**, Overall accuracy for different ALPaCA variants with various adaptors, including breakdowns for tumor-related (TCGA) and non-tumor-related (GTEx) multiple-choice and True/False questions. **c**, Performance lead-board of different ALPaCA variants across various question categories for TCGA and GTEx, respectively. **d**, Performance of ALPaCA-Hybrid across different cancer projects and tissue types, along with the trend line of accuracy. The bubble size represents the number of instructions from each cancer project or tissue type used in training, arranged from smallest to largest along the x-axis.

Except for the most basic Sampling and KMeans variants, all other ALPaCA variants achieved an overall accuracy of over 90%, with ALPaCA-Hybrid demonstrating the best overall performance at 91.76% (**Fig. 2b**). This variant performed best in tumor-related multiple-choice questions, where it achieved an accuracy of 91.20%, outperforming the second-best variant (ALPaCA-GMM) by 1.81%. It also achieved the highest accuracy in tumor-related True/False questions (89.05%) and non-tumor-related multiple-choice questions (96.60%). However, its performance was relatively weaker in non-tumor-related True/False questions (92.80%).

Further assessment of ALPaCA variants across different question categories (**Fig. 2c**) reveals that all models exhibited competitive performance for most non-tumor-related questions. For tumor-related questions, ALPaCA-Hybrid and ALPaCA-LongFormer achieved superior performance, consistently outperforming other variants across multiple categories. However, three specific categories of tumor-related questions exhibited relatively lower performance across all categories: tumor grading and staging, invasion characteristics, and tumor micro-environment (**Extended Data Fig. 2**). The primary challenge with these categories is that they not only require global slide-level information but also demand detailed insights from specific regions or information integration across multiple local regions. Such tasks necessitate the ability to guide the selection of visual tokens based on the question’s context. Consequently, ALPaCA-Hybrid and ALPaCA-LongFormer achieved better performance in these categories due to their vision-text interaction mechanisms. Although ALPaCA-QFormer incorporates vision-text interaction, its BERT-based architecture has significantly more parameters, requiring extensive pretraining data for effective initialization—beyond the scope of this study.

Additionally, we analysed the performance of the best-performing variant, ALPaCA-Hybrid, across different cancer projects and tissue sites to evaluate potential biases and determine whether such biases are correlated with the amount of training data (**Fig. 2d**). Our findings indicate that for the majority of cancer projects (24/28) and tissue sites (36/40), the accuracy exceeded 85%, with the lowest accuracy (TCGA-CHOL and Esophagus-Mucosa) still above 75%. Furthermore, as the amount of training data increased, ALPaCA’s performance across different cancer projects and tissue sites did not show significant variability, suggesting that its performance is primarily influenced by the difficulty of specific subsets of questions rather than by the training data size of individual subsets. For example, in the testset of TCGA-CHOL, out of 16 questions, there were 4 incorrect responses. Among these, 2 were related to invasion characteristics and 1 to staging, where pathologists reviewing the original slides confirmed that the results were difficult to determine based solely on the information available in the slides. The remaining error occurred in subtype classification, where the correct answer was “Cholangiocarcinoma”, but ALPaCA-Hybrid responded with “Hepatocellular carcinoma”. Upon review, pathologists noted that the histological features in the target slide were atypical, making it challenging to distinguish between these two subtypes. In the Esophagus-Mucosa test set, 5 out of 21 responses were incorrect. Notably, 4 of these errors were related to the same slide (GTEX-OHPK-0726), and pathologists confirmed that 3 of them were actually correct. This inconsistency stemmed from an error in the original pathology note, which stated that “there is hardly any mucosa in this section.” However, upon expert review, clear mucosal tissue was indeed present.

### 2.3 Automatic evaluation of open-ended questions

Beyond close-ended questions, we evaluated the performance of ALPaCA variants on the open-ended general pathology QA task, comprising 1,641 questions across tumor and non-tumor cases. This task better simulates real-world pathology diagnostics, where no predefined candidate options or even standard correct answers exist, making it crucial to evaluate whether ALPaCA can generate reasonable and relevant responses to the given questions and target slides. Given the challenges of evaluating openended QA, where correctness cannot be directly determined as in close-ended QA, we designed a comprehensive evaluation process (**Fig. 3a, Section 4.5**). In this section, we present the results of the automatic evaluation.

**Fig. 3:**
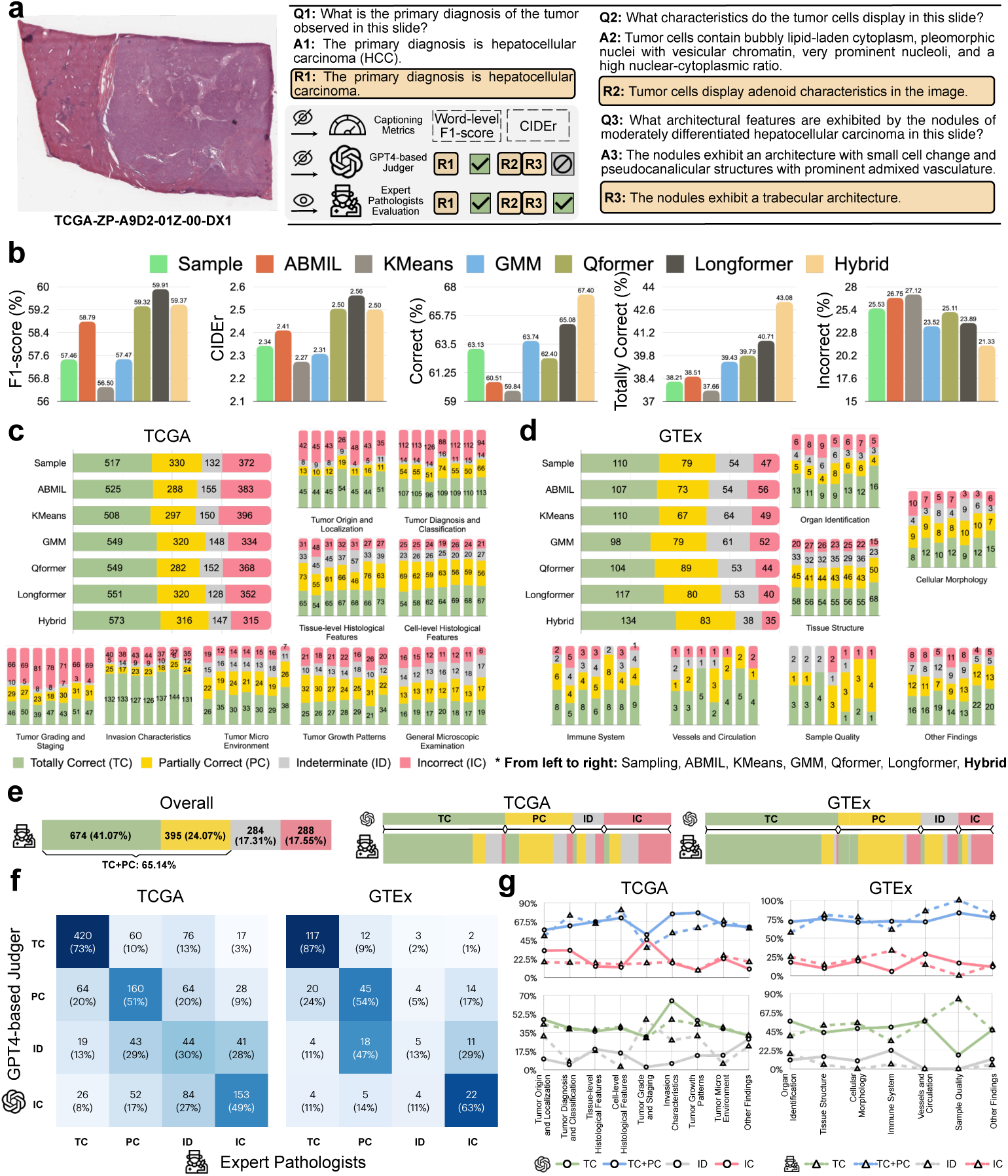
ALPaCA performance on open-ended questions. **a**, An example case of a liver tumor slide with corresponding open-ended questions, ground-truth answers, and ALPaCA-Hybrid’s responses, evaluated through three different assessment methods. **b**, Overall performance of different ALPaCA variants with various adaptors, measured by word-level F1-score, CIDEr, and correctness rates from the GPT-4-based judger. **c,d**, Detailed quantitative assessment results from the GPT-4-based judger for TCGA and GTEx, including breakdowns across different question categories. **e**, Overall performance of ALPaCA-Hybrid evaluated by expert pathologists, including their adjustments to GPT-4-based judger assessments. **f**, Comparison of expert pathologists’ assessments and GPT-4-based judger evaluations across four evaluation categories, and **g**, across question categories in TCGA and GTEx, respectively.

First, we calculated two commonly used automated evaluation metrics—word-level F1-score and CIDEr—to provide an initial assessment of response quality. Unsurprisingly, compared to fixed WSI token generation adaptors such as Sampling, KMeans, and GMM, ALPaCA variants with trainable adaptors (ABMIL, QFormer, Long-Former, and Hybrid) achieved superior results. Among them, ALPaCA-LongFormer performed the best, likely due to its ability to further refine patch embeddings, achieving 59.91% F1-score and 2.56 CIDEr (**Fig. 3b**). Overall, these trainable adaptors improve response quality by optimising how patch embeddings are aggregated, making the generated responses more “similar” to the ground truth answers. However, due to the nature of next-word prediction loss, this similarity is largely based on lexical overlap and word order rather than true semantic correctness. Thus, while higher word-level F1-score and CIDEr values indicate better alignment with the ground truth, they do not necessarily guarantee a qualitatively superior response.

Second, we used a custom GPT-4-based judger, which classified each response into one of four categories: “Totally Correct (TC)”, “Partially Correct (PC)”, “Incorrect (IC)”, or “Indeterminate (ID)”, based on its relevance to the question, consistency with the ground truth answer, and the reasonableness of the question itself (**Section 4.5.1**). Leveraging GPT-4’s strong comprehension abilities, this evaluation focused more on whether the response accurately conveyed the meaning of the ground truth answer rather than relying solely on word-level statistical overlap. From the overall performance results (**Fig. 3b**), we found that among the six adaptors, GMM and LongFormer demonstrated significant advantages. Both achieved an overall correct rate (the sum of TC and PC rates) of approximately 64%-65%. GMM exhibited a lower IC rate (23.52% vs. 23.89%), while LongFormer achieved a higher TC rate (40.71% vs. 39.43%). ALPaCA-Hybrid outperformed all other variants by a substantial margin. Its overall correct rate reached 67.40%, exceeding the second-best variant (ALPaCA-LongFormer) by 2.32%, while its TC rate (43.08%) surpassed ALPaCA-LongFormer by 2.37%. Additionally, its IC rate (21.33%) was 2.19% lower than that of the second-best variant (ALPaCA-GMM).

Moreover, we analysed the performance of different ALPaCA variants separately on tumor (TCGA) and non-tumor (GTEx) cases, along with their performance across corresponding question categories (**Fig. 3c,d**). In general, ALPaCA-GMM and ALPaCA-LongFormer performed competitively in tumor cases, while ALPaCA-GMM exhibited weaker performance in non-tumor cases. This can be attributed to the origin data of GTEx, where pathology notes are written for individual slides and provide relatively detailed histological descriptions, such as vascular stroma, fat, and fibrous tissue. These descriptions correspond to questions targeting specific regions within the slide, making vision-text interactive adaptors more advantageous. As expected, ALPaCA-Hybrid achieved the best results across both tumor and non-tumor cases. Examining performance across different question categories, ALPaCA-GMM excelled in tasks requiring global slide-level information, such as tumor origin and localization (TCGA), tumor diagnosis and classification (TCGA), and organ identification (GTEx). However, for questions demanding localized regional information or the integration of multiple regions, ALPaCA-LongFormer demonstrated superior performance. This advantage was particularly evident in tumor grading and staging (TCGA), cell-level histological features (TCGA), invasion characteristics (TCGA), cellular morphology (GTEx), immune system (GTEx), and vessels and circulation (GTEx). By combining these two adaptors, ALPaCA-Hybrid achieved competitive or superior results across all tasks. Notably, it demonstrated dominant performance in tumor micro-environment (TCGA) and tissue structure (GTEx) related tasks, further validating that integrating both global and local information enhances overall performance across diverse pathology tasks.

### 2.4 Pathologist evaluation of open-ended questions

To further validate the reliability and clinical relevance of ALPaCA, we present the expert evaluation results in this section. Two consultant pathologists manually assessed the responses generated by the best-performing variant, ALPaCA-Hybrid, as identified by the GPT-4-based judger. They followed the same evaluation criteria but had the additional capability to analyse the original slides, offering a more accurate and unbiased assessment (**Section 4.5.2**).

Overall, ALPaCA-Hybrid maintained consistent performance under expert pathologist evaluation, further confirming the effectiveness and reliability of the GPT-4-based judger as an automated evaluation method. Specifically, while the overall correct rate (TC+PC) decreased by 2.26%, the IC rate also dropped by 3.78%. This reduction was primarily due to expert pathologists classifying more tumor-related (TCGA) cases as ID—even when ALPaCA-Hybrid’s response matched the ground truth—stating that these questions could not be reliably answered based on the given slide alone (**Fig. 3e**). Additionally, expert pathologists and the GPT-4-based judger demonstrated higher consistency in non-tumor cases compared to tumor cases. This discrepancy stems from dataset differences—GTEx provides more precise slide descriptions, whereas

TCGA pathology reports contain greater noise. Across the four evaluation categories, expert pathologists and the GPT-4-based judger exhibited strong agreement in TC judgements, with 73% and 87% consistency in TCGA and GTEx cases, respectively. Notably, only 3% and 1% of cases were corrected to IC. The greatest divergence occurred in the ID category, where responses classified as ID by the GPT-4-based judger were reassigned in 70% and 82% of cases, primarily to PC or IC upon expert review (**Fig. 3f**). This discrepancy is closely related to the inherent limitations of LLMs, which lack the capability to observe WSIs directly. As a result, the GPT-4-based judger struggles to accurately classify ID questions.

Moreover, we observed variations in consistency between expert pathologists and the GPT-4-based judger across different question categories (**Fig. 3g**). For instance, in TCGA cases, tumor grading and staging, invasion characteristics, and tumor growth patterns, pathologists re-classified a significant portion of questions as ID, as they determined these questions could not be reliably answered based on the given slide alone. Surprisingly, 44 out of 94 responses in tumor diagnosis and classification were reclassified by expert pathologists from IC to PC or TC. These corrections stemmed from various reasons, all of which were considered correct by expert pathologists: some responses were diagnostically accurate but did not have the same level of granularity as the ground truth (e.g., the ground-truth answer was lung micropapillary carcinoma, while the response was lung adenocarcinoma). Others were influenced by updates in diagnostic guidelines, where certain rare subtypes were reclassified into broader categories (e.g., breast medullary carcinoma being categorized as infiltrating duct carcinoma). Additionally, some discrepancies arose because ground-truth answers were generated based on a patient’s ICD-O coding, whereas the actual clinical diagnosis differed (e.g., the ground-truth answer was teratocarcinoma, while the response was mixed germ cell tumor).

In GTEx cases, the highest inconsistencies were observed in the immune system and sample quality categories. For the immune system (18 Q&A pairs in total), 2 out of 3 ALPaCA-Hybrid responses initially classified as TC by the GPT-4-based judger were downgraded to PC by expert pathologists due to minor wording inaccuracies (e.g., lymphohistiocytic infiltrate vs. lymphoid infiltrate). Additionally, 2 out of 3 responses labeled as ID by the GPT-4-based judger were reclassified as incorrect after pathologists reviewed the slides. For the sample quality (6 Q&A pairs in total), 4 out of 5 responses were upgraded from IC or PC to TC. For example, in response to the question “What is the condition of the specimens seen in the image?”, the groundtruth answer provided a more detailed description (e.g., “Clean specimens with a good tube.”), whereas ALPaCA-Hybrid’s response (“The specimens are clean.”) still fully addressed the question. While the GPT-4-based judger classified it as PC due to missing details, expert pathologists confirmed it as TC, recognizing it as an accurate and complete response.

Additionally, expert pathologists’ evaluation of the open-ended testset curated a subset of answerable questions for “General Slide-QA” dataset. We then re-evaluated all ALPaCA variants on this expert-curated testset using the automatic evaluation method to verify potential biases in the initial assessment in Section 2.3. We found that the conclusions remained basically unchanged, with ALPaCA-Hybrid’s advantage even slightly increasing (**Extended Data Fig. 5**). Compared to the second-best ALPaCA variant, its overall correct rate advantage is by an additional 1.36%, while its TC rate advantage increased by 1.17%. More detailed analyses on open-ended performance across different cancer projects and tissue sites, as well as assessments by joint or individual expert pathologists, are presented in **Extended Data Fig. 3** and **4**.

### 2.5 Accommodating in-depth disease-specific question answering

Finally, we evaluated the performance of ALPaCA-Hybrid after fine-tuning on two disease-specific QA datasets: “Specific Slide-QA (Breast)” and “Specific Slide-QA (Morphology)”. The former focuses on detailed molecular and morphological aspects of breast cancer, including molecular subtypes, lymphovascular invasion, necrosis, nuclear pleomorphism, fibrous focus, mitosis, and other key pathological features. (**Fig. 4a**) The latter encompasses fine-grained histological morphology QA for gastric cancer, clear cell renal carcinoma, and ovarian cancer. (**Fig. 4b**)

**Fig. 4:**
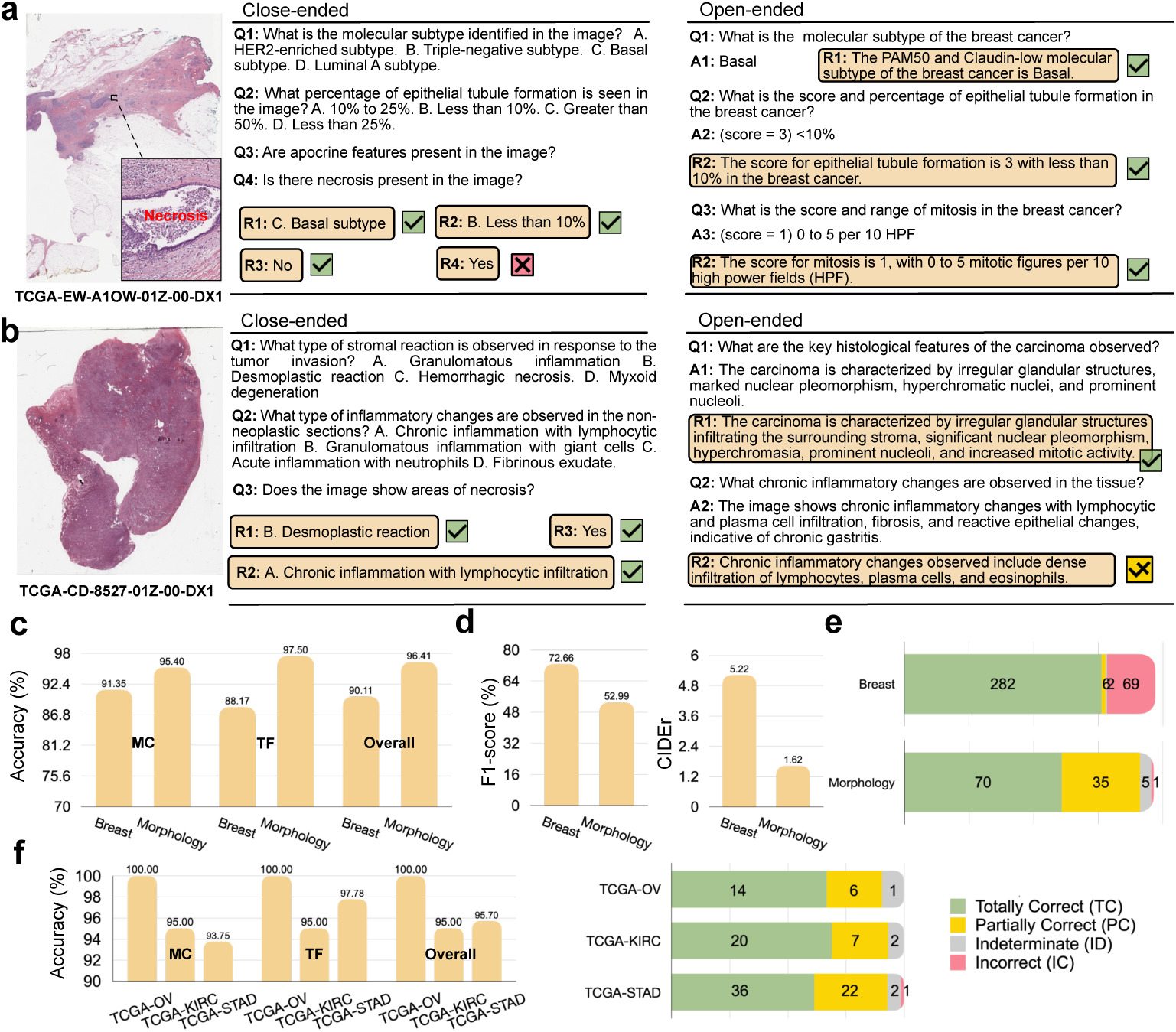
ALPaCA performance on specific QA datasets. **a,b**, Example cases of a breast tumor slide from the “Specific Slide-QA (Breast)” dataset and a gastric tumor slide from the “Specific Slide-QA (Morphology)” dataset, with corresponding close- and open-ended questions, ground-truth answers, and ALPaCA-Hybrid’s responses, evaluated by the GPT-4-based judger. **c**, Close-ended performance of ALPaCA-Hybrid on the two specific datasets, including overall accuracy, as well as accuracies for multiple-choice and true/false questions separately. **d,e**, Open-ended performance of ALPaCA-Hybrid on the two specific datasets, including captioning metrics and detailed quantitative assessment results from the GPT-4-based judger. **f**, Performance of ALPaCA-Hybrid across three cancer projects in the “Specific Slide-QA (Morphology)” dataset.

Overall, ALPaCA-Hybrid achieved performances on the two disease-specific QA datasets that were comparable to or even exceeded its results on the general QA dataset. Specifically, in the “Specific Slide-QA (Breast)” test set, the close-ended accuracy was 90.11%, slightly lower than the 96.41% achieved on “Specific Slide-QA (Morphology)” (**Fig. 4c**). This discrepancy is attributed to the higher difficulty of questions in “Specific Slide-QA (Breast)”, which includes not only molecular subtypes but also quantitative morphological questions. In contrast, while “Specific Slide-QA (Morphology)” focuses on more fine-grained morphological details, most questions involve identifying whether a particular morphological feature is present or determining its classification, making the close-ended task relatively easier.

For open-ended performance, ALPaCA-Hybrid significantly outperformed in “Specific Slide-QA (Breast)”, achieving a word-level F1-score of 72.66% vs. 52.99% and a CIDEr score of 5.22 vs. 1.62 compared to “Specific Slide-QA (Morphology)” (**Fig. 4d**). This is because “Specific Slide-QA (Breast)” open-ended questions were generated from structured tabular data, resulting in simpler, more standardized responses that the model could fully reproduce. In contrast, the “Specific Slide-QA (Morphology)” dataset contains free-text descriptions with higher variability, making it more challenging for the model to generate responses that exactly match the ground truth. However, based on evaluations by the GPT-4-based judger (**Fig. 4e**), even when ALPaCA-Hybrid’s responses did not precisely match the correct answers, they often conveyed similar meanings or contained the correct information. In “Specific Slide-QA (Morphology)”, ALPaCA correctly answered 105 out of 111 open-ended questions (70 were TC) and made only one IC response. In “Specific Slide-QA (Breast)”, ALPaCA-Hybrid correctly answered 288 out of 359 open-ended questions, with 282 TC, achieving an accuracy of over 80%.

Additionally, as shown in **Fig. 4a**, annotation errors exist in certain breast cancer cases. The slide was initially labelled as necrosis absent, whereas ALPaCA-Hybrid detected its presence. Upon further review, pathologists confirmed small necrosis areas in the central region of the slide, underscoring ALPaCA’s capability to identify overlooked pathological details.

## 3 Discussion

In this study, we have introduced ALPaCA, a slide-level LVLM designed for WSI question answering. We demonstrated that connecting a foundation pathology vision encoder with an LLM via specialized adaptors enables effective slide-level QA. We also curated a large-scale WSI-level QA dataset spanning multiple cancer types and tissue sites, providing a foundation for developing LVLMs for WSI analysis. This demonstrated the feasibility of automatically constructing extensive WSI-level QA datasets. Additionally, we designed a comprehensive evaluation framework for slide-level LVLMs, ranging from simple close-ended question accuracy to automated scoring of open-ended responses and expert pathologist assessments. After a detailed comparison of six candidate adaptors, we proposed the hybrid adaptor (ALPaCA-Hybrid), which achieved the best performance across multiple evaluation scenarios. Furthermore, we showed that fine-tuning ALPaCA on specific datasets enables in-depth, disease-specific interactive QA, supporting more complex and specialized pathology diagnostic workflows.

Despite these advancements, this work has certain limitations. First, although we curated the largest publicly available dataset for this task, the overall data volume remains relatively limited, especially for rare tumors and real-world non-tumor cases. The GTEx database, primarily composed of postmortem donor tissues, differs from suspected tumor slides or adjacent normal tissue sections commonly encountered in actual diagnostic settings. Second, the experimental analyses of this study, particularly expert pathologist evaluations, revealed the challenges of generating slide-level descriptions and QA pairs from patient-level pathology reports. Since a single pathology report often encompasses information from multiple slides, it inevitably results in questions that cannot be reliably answered using just one representative slide, as observed in the TCGA portion of the “General Slide-QA” dataset. However, the GTEx portion demonstrated that when descriptions are curated specifically for individual slides, current LLMs can automatically generate high-quality slide-level QA datasets with minimal ambiguity. This highlights two promising directions for future slide-level LVLM research: improving dataset quality by collecting fine-grained, per-slide textual descriptions or developing multi-slide integration frameworks for patient-level question answering. Regardless of the approach, ALPaCA serves as a flexible foundation for integrating both strategies, advancing more precise and context-aware pathology QA. Finally, ALPaCA lacks explicit spatial grounding and deep reasoning capabilities, limiting both interpretability and diagnostic reliability. It cannot pinpoint specific slide regions associated with its answers, which is particularly challenging for quantitative tissue or cellular analysis tasks requiring precise localization and measurement—an issue that could be addressed by integrating dedicated detection and segmentation models. Additionally, as an instruction-tuned model, ALPaCA does not yet leverage reinforcement learning-based decision-making, which could further enhance its reasoning abilities and overall clinical applicability.

In conclusion, ALPaCA, as a slide-level LVLM, holds significant potential for both research and clinical applications. With further refinement and validation on real-world datasets, ALPaCA can evolve into a more robust and clinically applicable slide-level LVLM. It has the potential to drive large-scale pathological data mining in cancer research while enhancing automated pathology reporting, diagnostic decision-making, and second-opinion validation in clinical practice. Furthermore, ALPaCA’s modular framework ensures adaptability to future advancements. Both the vision encoder and LLM backbone can be updated with the latest models as research progresses, enhancing performance and generalisation. Beyond its direct applications, ALPaCA’s framework, training strategy, and evaluation process provide a structured foundation for future advancements in slide-level LVLMs, contributing to the development of more robust and clinically relevant AI-driven WSI analysis solutions.

## 4 Methods

### 4.1 Curation of datasets

First, we curated a slide-level caption dataset, “General Slide-Caption”, comprising 35,913 WSIs with corresponding descriptions. The source data was derived from the two largest publicly available human tissue databases: TCGA, which includes 32 cancer studies and 134 diagnosis types, and GTEx, which spans 40 non-tumor tissue sites. For TCGA, each WSI (FFPE diagnostic slide stained with H&E at 40x or 20x magnification) is paired with a patient-level pathology report (in PDF format) and basic patient metadata. The Python package “PyMuPDF” converted PDF files into text files. For PDFs in image format, which could not be processed automatically, optical character recognition (OCR) techniques were applied. The raw pathology reports often included irrelevant sections, such as gross examination. We used the commercial solution ChatGPT (GPT-3.5 Turbo) to summarize each raw report into a structured text description. This process excluded any gross examination information while retaining all microscopic pathological findings. Next, we refined these descriptions using the state-of-the-art general-purpose version of ChatGPT (GPT-4o) with patient metadata and well-designed prompts. This step ensured that the descriptions were accurate and tailored to specific patient contexts, focusing exclusively on the content of primary tumor slides, the main type of TCGA diagnostic slides. For GTEx, each WSI (FFPE diagnostic slide stained with H&E at 20x magnification) is accompanied by a pathology note and basic tissue metadata. Since the pathology notes are well-structured and specific to the corresponding WSI, we only rephrased them using GPT-4o with the aid of tissue metadata and carefully designed prompts. This process excluded any quantitative tissue information (*e.g.*, length, thickness) that is hard to infer from WSIs.

Second, we curated a slide-level QA dataset, “General Slide-QA”, comprising 341,051 instructions. This dataset includes True or False and Multiple-Choice closed-ended Q&A pairs, along with open-ended pairs requiring free-text responses. Based on the “General Slide-Caption” dataset, slide-level Q&A pairs were generated by GPT- 4o using prompts with clear guidelines, designed by experienced pathologists, tailored for close-ended and open-ended scenarios of tumor and non-tumor cases, respectively.

Lastly, two specific and in-depth slide-level QA datasets were curated: “Specific Slide-QA (Breast)”, focusing on TCGA-BRCA cases, and “Specific Slide-QA (Morphology)”, covering TCGA-STAD, TCGA-KIRC, and TCGA-OV cases. Instead of utilizing the general descriptions generated for the “General Slide-Caption” dataset, we incorporated additional molecular annotations of TCGA-BRCA cases, as provided in [42], to create an in-depth molecular QA dataset. Furthermore, we leveraged the detailed morphological descriptions generated by PathChat, the state-of-the-art pathology AI copilot [36], to construct the in-depth morphology QA dataset. Specifically, for each WSI, we extracted 1024 ⇥ 1024 patches at 40⇥, 20⇥, and 10⇥ magnifications and applied KMeans clustering to obtain 32, 16, and 8 representative clusters, respectively. PathChat was then used to generate morphological descriptions for the patches closest to each cluster center, and these descriptions were subsequently summarized into comprehensive slide-level reports by GPT-4o. Using these enriched annotations and descriptions, we curated specific, detailed, close-ended, and open-ended Q&A pairs tailored to breast, gastric, renal, and ovarian cancers.

For testing and evaluation, we randomly selected 3 cases per non-tumor tissue site from the GTEx database and 1 or 2 cases per diagnosis type from the TCGA database as hold-out test cases, totaling 337 cases, with the remaining cases used for training. Notably, these hold-out cases were excluded from all stages of training to prevent data leakage, ensuring an unbiased evaluation. This split was applied consistently across all three curated datasets. Details of the source data are presented in **Extended Data Table 1**, while information on the curated datasets and data splits is provided in **Extended Data Table 2**. Additional statistical insights into the “General Slide-QA” dataset are illustrated in **Extended Data Fig. 1-2** and **Extended Data Table 3-6**. All prompts used for data curation are detailed in **Supplementary information**.

### 4.2 Pre-processing of WSIs

Based on the general pre-processing pipeline for WSI analysis, we began by detecting tissue regions in each WSI and dividing them into small patches with a 50% overlap. Since different questions for the same case or the same question across different cases may require information from varying levels of the WSI or their combination, WSI tessellation was performed hierarchically. Patch sizes were set to 1024, 2048, and 4096 at 40x magnification. If the maximum magnification of the original slide was 20x, the patch sizes were adjusted to 512, 1024, and 2048, respectively, to maintain consistency. These patches were then resized to a uniform size (224 ⇥ 224), mimicking tiles at 40x, 20x, and 10x magnifications to capture information across multiple levels of each WSI. Then, we used the frozen CONCH [41] to extract features from patches at different magnifications, forming hierarchical patch embeddings. CONCH, a state-of-the-art pathology image encoder, is aligned with text using 1.17 million pathology image-caption pairs. CONCH is better suited for LLM integration than other pre-trained vision-only pathology image encoders. Furthermore, as the image-text alignment data is not restricted to specific magnifications, it is particularly effective for extracting patch embeddings across various magnification levels.

### 4.3 ALPaCA architecture and various adaptors

Built upon the hierarchical patch embeddings of each WSI, it is necessary to compress and aggregate the patch embeddings into corresponding WSI tokens for two primary reasons: (1) the raw patch embeddings are too sparse, containing redundant information, and (2) the sequence length of uncompressed patch embeddings would be excessively long, making it impractical for LLM training. To achieve this, six distinct types of adaptors were proposed and implemented, designed to aggregate patch embeddings at different magnifications into WSI tokens. These tokens were then concatenated with text tokens in a specific structure for LLM training. Specifically, we adopted the 8-billion-parameter version of Llama3.1, a decoder-only transformer-based autoregressive language model, as the LLM for ALPaCA due to its state-of-the-art performance, accessibility as a publicly available model, and manageable training cost, making it suitable for academic research. We implemented four vision-only adaptors and two vision-text interactive adaptors. The key difference lies in whether the adaptors incorporate any text context, such as questions, during the compression and aggregation of WSI tokens. The detailed descriptions of each adaptor are as follows:

- **Sampling:** The most straightforward approach involves sampling a subset of patch embeddings to represent the WSI. To ensure the representativeness of the sampling, patch embeddings are grouped into clusters using the KMeans algorithm, and one patch embedding is randomly sampled from each cluster to serve as a token in the final WSI representation.
- **KMeans:** Another straightforward method involves clustering the patch embeddings and using the centroid of each cluster as a token in the WSI representation. This approach ensures that each token represents the central tendency of the corresponding cluster, providing a compact and representative summary of the WSI.
- **GMM [43]:** A prototype-based approach rooted in the Gaussian Mixture Model (GMM) is used to summarize patch embeddings into a set of morphological prototypes. Specifically, each patch embedding is assumed to be generated from a mixture distribution, where each mixture component represents a morphological exemplar. The resulting prototypes provide a compact representation of the WSI by capturing the dominant patterns present in patches.
- **ABMIL [44]:** A classic aggregation method in WSI analysis, Attention-Based Multi-Instance Learning (ABMIL) uses a multi-head gated attention network to aggregate patch embeddings into diverse WSI tokens. This method dynamically assigns attention weights to patches, ensuring that the features most relevant to correct answers contribute most significantly to the final WSI representation.
- **Qformer [27]**: A state-of-the-art vision-text interaction adaptor, that incorporates both self-attention and cross-attention layers. It begins by initializing multiple learnable query tokens, which are first refined through a self-attention mechanism applied to the query tokens and question embeddings (from the LLM tokenizer). The optimized query tokens are then used in the cross-attention layers to query the patch embeddings, enabling question-specific refinement of the query tokens for the downstream QA task. To simplify the adaptor, we utilized a lightweight design for each magnification level. Specifically, a BERT encoder with a single hidden layer and a single cross-attention layer was employed.
- **LongFormer**: A custom-designed vision-text interaction adaptor, LongFormer differs from QFormer by not merely querying fixed patch embeddings for task-specific WSI representations but also enabling the optimization of patch embeddings. By utilizing a LongNet structure [45], it efficiently handles ultra-long sequence feature optimization, allowing for better integration and refinement of information across the entire WSI. This design ensures that patch embeddings dynamically adapt to the specific task requirements, enabling the generation of more precise and context-aware WSI tokens.

All adaptors produced 32, 16, and 8 WSI tokens for 40x, 20x, and 10x magnifications, respectively. A resampler (composed of one hidden layer, ReLU activation, and a dropout rate of 0.25) then maps these WSI tokens to the required 4096 dimensions for Llama3.1.

Except for the GMM adaptor, which produces 1025-dimensional tokens, all other adaptors generate 512-dimensional tokens, consistent with the original patch embedding dimensions. Note that Sampling, KMeans, and GMM adaptors do not include trainable parameters aside from the resampler. ABMIL uses a trainable gated attention network with three linear layers and 256 hidden units. QFormer and LongFormer have a reducer with two low-rank hidden layers that map question tokens from 4096 to 512 dimensions: the first reduces the dimension from 4096 to 128, and the second further reduces it to 512, followed by a ReLU activation. The cross-attention part for both of them is implemented using a single “MultiheadAttention” layer with 512 hidden states and 8 heads. For the self-attention part, QFormer employs a BertModel with one hidden layer, 512 hidden states, and 8 heads. LongFormer uses a regular transformer block with one self-attention layer (512 hidden states, 8 heads) and one feed-forward layer (2048 hidden states). Additionally, LongFormer integrates a two-layer LongNet with 512 hidden states and 8 heads to optimize patch embeddings concurrently.

Moreover, due to the outstanding performance of GMM and LongFormer adaptors in answering different types of questions, the best-performing ALPaCA variant (**ALPaCA-Hybrid**) is derived from their combination. Specifically, the WSI tokens extracted by both adaptors are combined additively after the resampler layer and then fed into Llama3.1.

### 4.4 Three-stage training of ALPaCA

For training, we adopted a three-stage strategy, with the first two stages commonly used in LVLM training. The first stage, termed “**Comprehension**”, involves freezing the LLM weights while updating only the adaptors. This stage is supervised, with ALPaCA trained to predict the description corresponding to each WSI in the “General Slide-Caption” dataset. The adaptors learn to aggregate patch tokens into WSI tokens and project the image space into the shared embedding space of text tokens utilized by the LLM. The second stage, termed “**Instruction**”, involves training both the LLM and adaptors end-to-end. In this stage, ALPaCA is trained to generate responses to diverse general instructions from the “General Slide-QA” dataset. The third stage, termed “**Specialisation**”, fine-tunes both the LLM and adaptors end-to-end on specific and in-depth datasets. In this stage, ALPaCA is fine-tuned to address domain-specific questions related to a particular tissue or cancer type. For this, we use two “Specific Slide-QA” datasets, one focusing on the molecular aspects of breast cancer and the other on the fine-grained morphological patterns of gastric, renal, and ovarian cancers.

Specifically, we use the next-word prediction loss across all three stages, treating ALPaCA as an autoregressive language model. The loss is calculated only for the answer text tokens, and the prompt is included solely for close-ended instructions. The structure of the LLM input list is as follows:

<|Image|> <|H|> 40x WSI tokens <|M|> 20x WSI tokens <|L|> 10x WSI tokens <|Question|> question text tokens <|Prompt|> prompt text tokens <|Answer|> answer text tokens <|pad|> <|pad|> <|pad|>

Note that the first stage also follows this input structure, transforming the captioning task into a QA task by setting several fixed questions to guide the caption generation. The learning rate was set to 2.0e-5, with a gradient accumulation step size of 8. The training batch size was 4, and the warmup phase included 1000 steps. The training process spanned 20 epochs for the first stage and 5 epochs for the second and third stages. The total training time amounted to approximately 60 GPU hours for Stage 1, 634 GPU hours for Stage 2, and 50.8 and 28.5 GPU hours for Stage 3 on the Specific Slide-QA (Breast) and Specific Slide-QA (Morphology) datasets, respectively.

### 4.5 ALPaCA evaluation

We compared different versions of ALPaCA, equipped with various adaptors or trained using different strategies. For close-ended questions, all versions of ALPaCA generated predicted choices in a consistent format (*e.g.*, ‘Yes’, ‘A. Grade III’), enabling direct calculation of evaluation metrics such as accuracy. For open-ended questions, we computed the word-level macro F1-score and the CIDEr score against the ground truth answers, with the CIDEr score being commonly used in captioning tasks. However, these automatic quantitative metrics often fail to reflect the quality of predictions accurately. To address this, we conducted further evaluations using a prompt-guided GPT-4o judger alongside manual assessments by professional pathologists.

#### 4.5.1 Evaluating with GPT-4o

We began by designing detailed assessment guidelines for GPT-4o, as outlined in the “Prompts” section of Supplementary. Since no commercial models currently support direct WSI input, we provided GPT-4o with the question, the corresponding ground truth answer, and ALPaCA’s response for evaluation. To minimize the randomness of GPT-4o evaluations, each response was assessed five times, and the final evaluation result was determined by taking the majority vote across the five evaluations. GPT- 4o primarily assessed the degree of alignment between these elements and categorized each response into one of four options: totally correct, partially correct, indeterminate, or incorrect, considering both the consistency between the ground truth answer and ALPaCA’s response, as well as the content of the question itself. It is worth noting that even though the ground truth answers were derived from pathology reports or notes, these reports are typically written for a collection of slides from each patient and can sometimes be overly general. As a result, the ground truth may lack completeness or fail to align perfectly with the single WSI observed by ALPaCA. Moreover, for open-ended questions, multiple valid answers often exist that may be correct but do not overlap entirely. These factors highlight the limitations of evaluations solely based on the alignment between the ground truth and the model’s response without directly examining the corresponding WSIs.

#### 4.5.2 Evaluating with Expert Pathologists

With access to the original WSIs, pathology reports/notes, and patient metadata, two consultant pathologists evaluated the best variant (Longnet) of ALPaCA’s responses to open-ended questions. To ensure objectivity and consistency, the evaluation process was conducted as follows:

- **Step 1:** Joint evaluation of a subset of cases (one case per tissue or tumor type). This step aimed to establish a consistent evaluation standard and ensure alignment between the two pathologists.
- **Step 2:** Independent evaluation of the remaining cases by each pathologist. This increased the efficiency of the evaluation process.
- **Step 3:** Joint discussion for cases that were difficult to determine. This step resolved ambiguities and ensured consensus on challenging cases.

In alignment with the GPT-4o evaluation process, each response was categorized into one of four options: totally correct, partially correct, indeterminate, or incorrect. Since they had access to the original WSIs, their evaluations were based on both the questions and the exact slides analyzed by the model rather than solely on answers extracted from reports or notes. This significantly enhanced the reliability of the assessments. Additionally, the GPT-4o evaluation results were provided for their reference.

### 4.6 Hardware and software

Python (v3.10.4) was used for all experiments and analyses in this study. All three stages of ALPaCA were trained on two 80 GB NVIDIA A100 GPUs configured for distributed training, utilizing the PyTorch (v2.2.1) deep learning framework with CUDA 11.8. Training acceleration was achieved through the integration of Accelerate (v0.31.0) with DeepSpeed (v0.14.0).

## 5 Data availability

The TCGA WSIs, pathology reports, and associated clinical metadata are accessible via the NIH Genomic Data Commons (https://portal.gdc.cancer.gov). The GTEx WSIs, pathology notes, and associated clinical metadata are available from the Genotype-Tissue Expression project (https://www.gtexportal.org/). The curated datasets will be publicly released on Hugging Face for research purposes, including: “General Slide-Caption” (CNX-PathLLM/TCGA-WSI-Description-4onew, CNX-PathLLM/TCGA-WSI-Description-4omini, CNX-PathLLM/GTEx-WSI-Description), “General Slide-QA” (CNX-PathLLM/TCGA-WSI-CloseQA-Balanced, CNX-PathLLM/GTEx-WSI-CloseQA-Balanced, CNX-PathLLM/TCGA-WSI-OpenQA, CNX-PathLLM/GTEx-WSI-OpenQA), “Specific Slide-QA (Breast)” (CNX-PathLLM/TCGA-BRCA-Details-CloseQA, CNX-PathLLM/TCGA-BRCA-Details-OpenQA), and “Specific Slide-QA (Morphology)” (CNX-PathLLM/PathChat-CloseQA-Balanced, CNX-PathLLM/PathChat-OpenQA).

## 6 Code availability

The code for training and evaluating ALPaCA will be released at https://huggingface.co/CNX-PathLLM/Llama-slideQA, along with the checkpoint for the best-performing variant, ALPaCA-Hybrid (Llama-slideQA.bin), and its fine-tuned versions on two specific slide-QA datasets (Llama-slideQA-BRCA.bin, Llama-slideQA-morphology.bin). Comprehensive documentation of the model architecture, training procedure, and evaluation protocols will also be provided to ensure full reproducibility for the research community.

## Supporting information

prompt for gpt generation

## Data Availability

All data produced in the present work are contained in the manuscript

---

**Extended Data Table 1:**
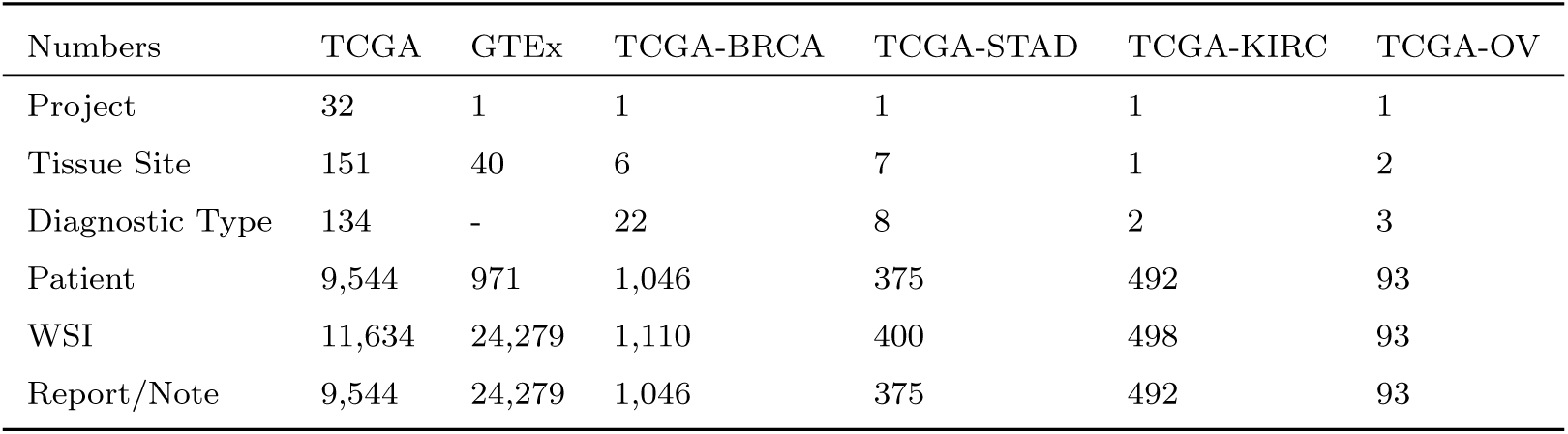
Statistics of Source Data.

**Extended Data Table 2:** Statistics of Curated Datasets.

| Dataset | Data Source | Question Type | Number of Q&A Pairs |  |  |
| --- | --- | --- | --- | --- | --- |
|  |  |  | Train | Test | Total |
| General Slide-Caption | TCGA | Description | 11,417 | 217 | 11,634 |
|  | GTE<sub>x</sub> | Description | 24,159 | 120 | 24,279 |
| General Slide-QA | TCGA | Open-ended | 60,441 | 1,351 | 61,792 |
|  | TCGA | Close-ended | 79,670 | 1,467 | 81,137 |
|  | GTE<sub>x</sub> | Open-ended | 47,017 | 290 | 47,307 |
|  | GTE<sub>x</sub> | Close-ended | 149,879 | 936 | 150,815 |
| Specific Slide-QA<br>(Breast) | TCGA-BRCA | Open-ended | 12,830 | 359 | 13,189 |
|  | TCGA-BRCA | Close-ended | 17,000 | 475 | 17,475 |
| Specific Slide-QA<br>(Morphology) | TCGA-STAD | Open-ended | 2,676 | 61 | 2,737 |
|  | TCGA-STAD | Close-ended | 3,886 | 93 | 3,979 |
|  | TCGA-KIRC | Open-ended | 3,694 | 29 | 3,723 |
|  | TCGA-KIRC | Close-ended | 5,102 | 40 | 5,142 |
|  | TCGA-OV | Open-ended | 581 | 21 | 602 |
|  | TCGA-OV | Close-ended | 916 | 34 | 950 |

**Extended Data Table 3:**
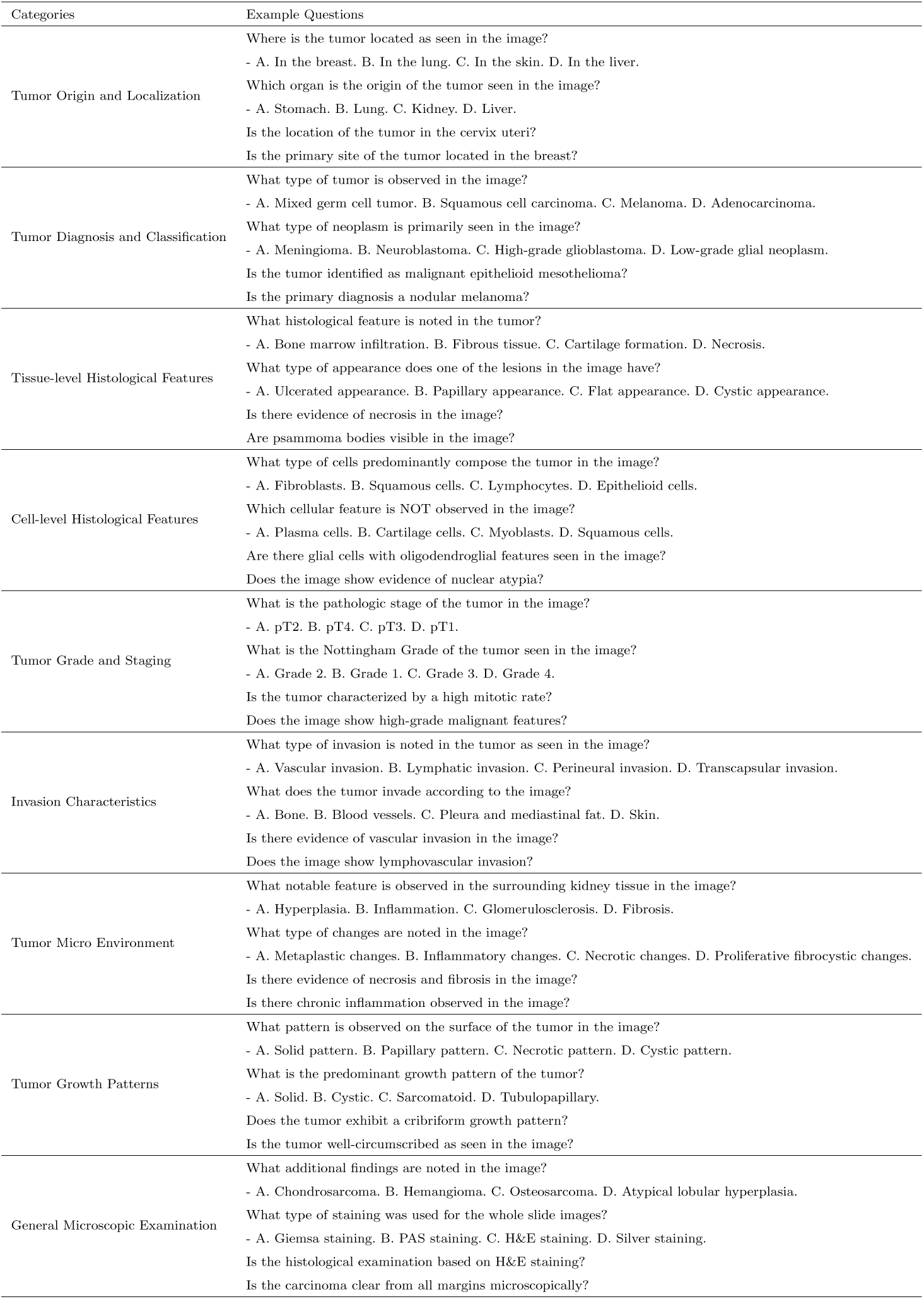
Example close-ended questions of different categories in “General Slide QA” dataset (TCGA).

**Extended Data Table 4:**
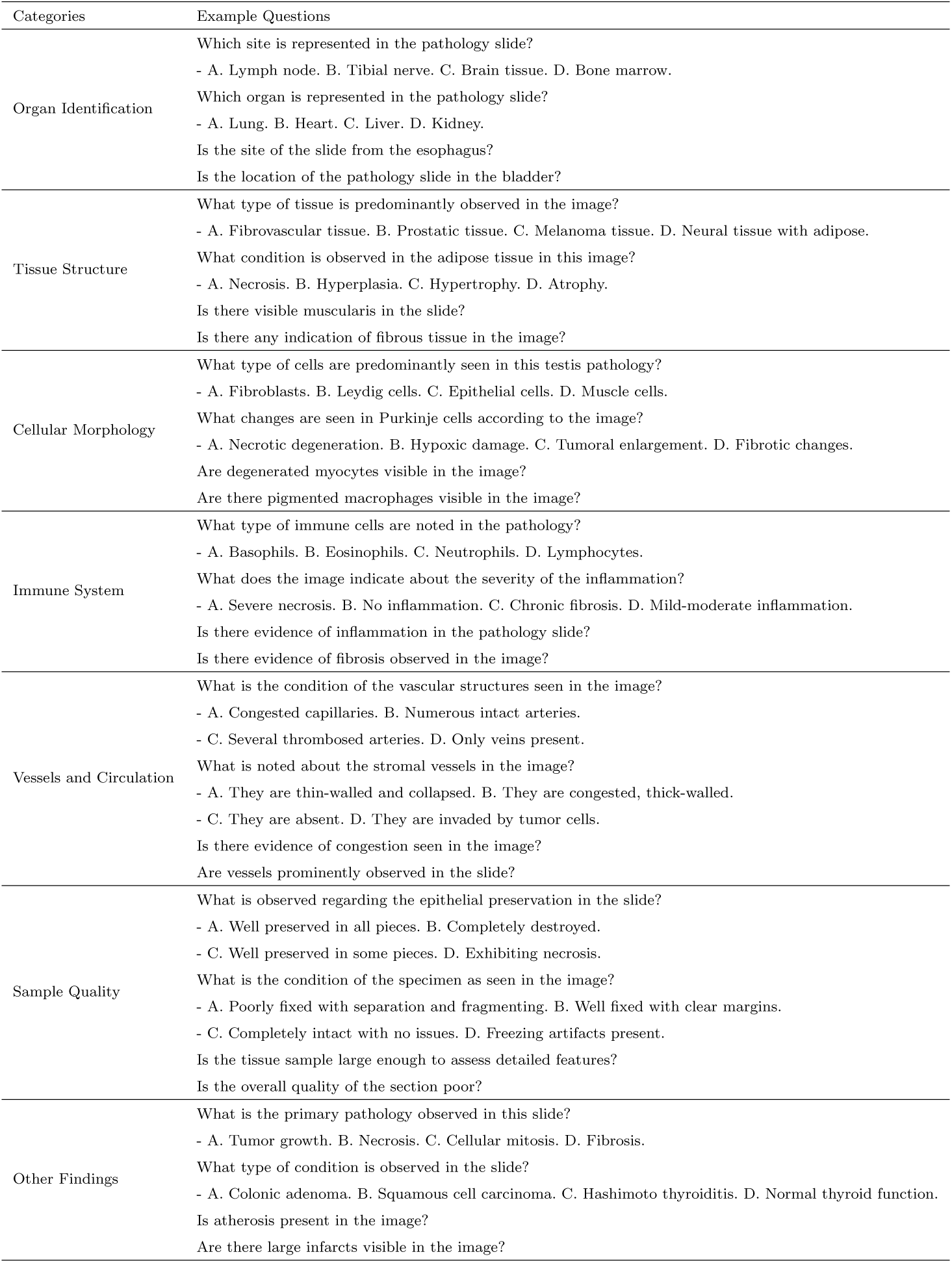
Example close-ended questions of different categories in “General Slide QA” dataset (GTEx).

**Extended Data Table 5:** Example open-ended questions of different categories in “General Slide QA” dataset (TCGA).

| Categories | Example Questions |
| --- | --- |
| Tumor Origin and Localization | Where is the tumor present microscopically in this slide? |
|  | Where does the tumor originate from according to this slide? |
|  | What is the original site of the tumor as seen in this slide? |
|  | From which tissues does the tumor in this slide originate? |
|  | From which part of the lung does the carcinoma in this slide originate? |
| Tumor Diagnosis and Classification | What type of tumor is identified in this slide? |
|  | What is the primary diagnosis of the tumor described in this slide? |
|  | How is the tumor diagnosed in terms of its histological type? |
|  | What subtype of carcinoma is mentioned in this slide? |
|  | How is the carcinoma specifically classified in this slide? |
| Tissue-level Histological Features | What is the histological composition of the tumor? |
|  | How does the tumor appear histologically in this slide? |
|  | Are there any areas of necrosis in the tumor as seen in the slide? |
|  | What histological features are observed in the primary tumor’s H&E slide? |
|  | What additional findings are present within the myometrium in the image? |
| Cell-level Histological Features | What nuclear features are observed in the cells in this slide? |
|  | What is the mitotic activity observed in this slide? |
|  | What cellular features are identified in this slide? |
|  | What are the characteristics of the epithelial cells seen in the image? |
|  | Does the tumor in this slide exhibit any mitotic activity? |
| Tumor Grade and Staging | What is the stage classification of this slide? |
|  | What histological grade is the tumor in this slide? |
|  | What is the nuclear grade of the tumor in this slide? |
|  | What histologic grade is assigned to the tumor seen in this slide? |
|  | What FIGO grade is the tumor classified as? |
| Invasion Characteristics | Which structures does the tumor invade as seen in this slide? |
|  | What tissues does the tumor invade in this slide? |
|  | What layers are invaded by the tumor as seen in this slide? |
|  | Is perineural invasion present in the melanoma as seen in this slide? |
|  | Does the tumor show any invasion in this slide? |
| Tumor Micro Environment | What type of reaction is observed around the tumor in this slide? |
|  | What kind of inflammatory response is induced by the tumor in this slide? |
|  | What is the level of lymphocytic infiltrate in the tumor in this slide? |
|  | What is found in the surrounding tissues of the tumor in this slide? |
|  | What evidence of tissue reaction is seen in this slide? |
| Tumor Growth Patterns | What growth patterns are observed in the tumor in this slide? |
|  | What histological growth pattern is observed in this slide? |
|  | What pattern do the tumor cells exhibit in this slide? |
|  | What components make up the tumor seen in this slide? |
|  | What is the composition of the stroma in the tumor? |
| General Microscopic Examination | What notable features are seen in this slide? |
|  | What evidence of pathology is shown in this slide? |
|  | What components does this slide consist of? |
|  | What staining technique is used in the slide of the tumor? |
|  | What additional findings are present in this slide? |

**Extended Data Table 6:**
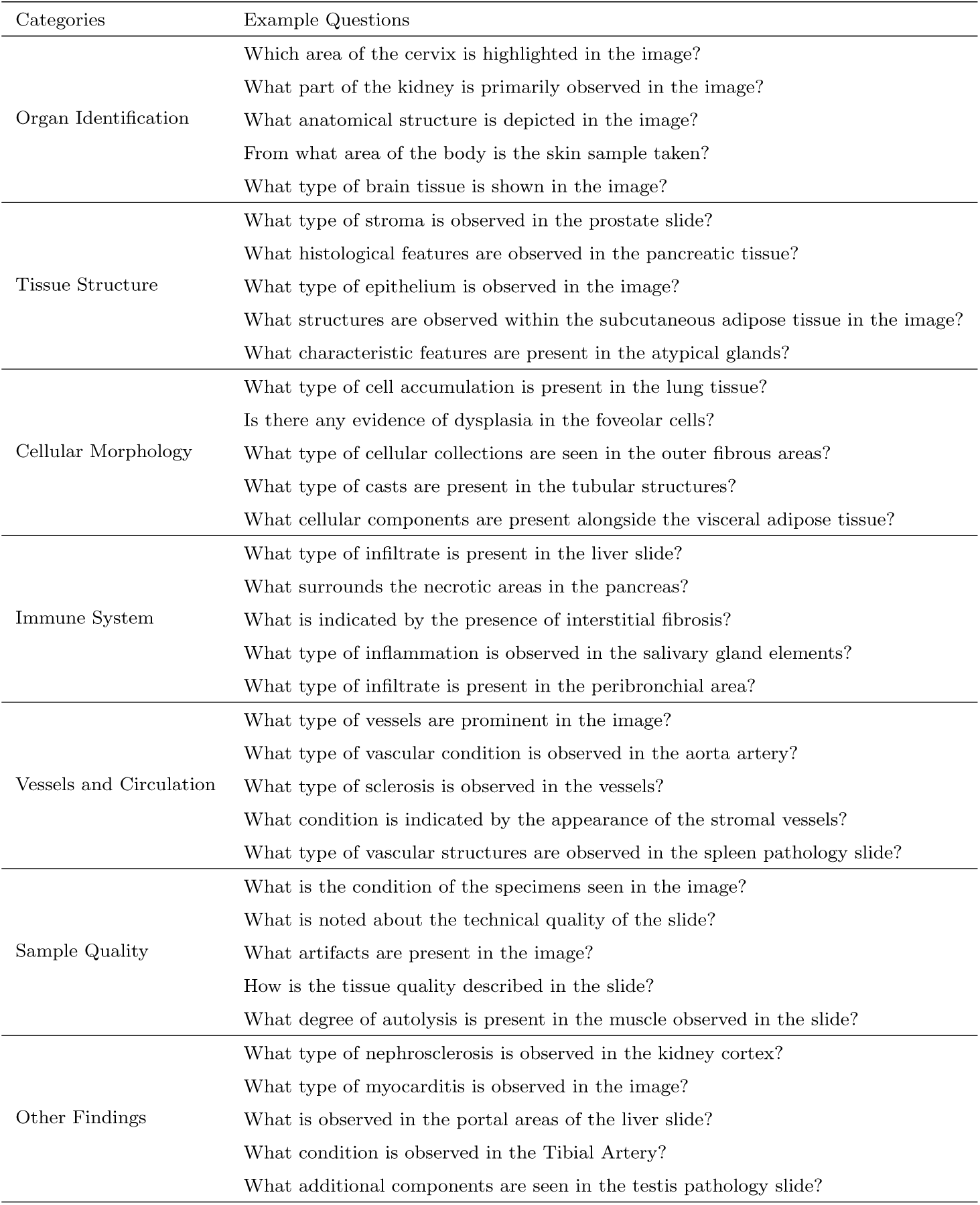
Example open-ended questions of different categories in “General Slide QA” dataset (GTEx).

**Extended Data Table 7:**
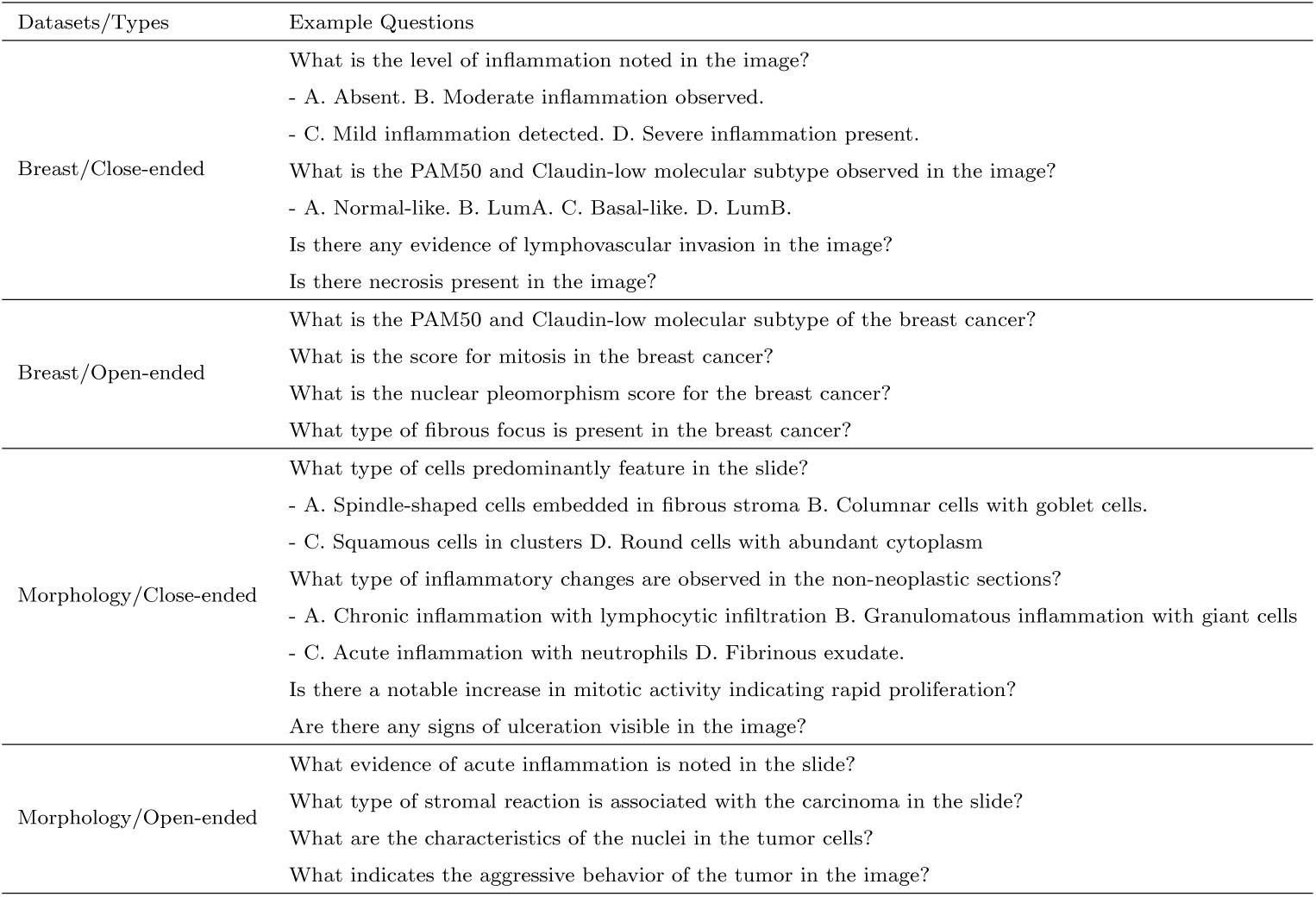
Example questions of two “Specific Slide QA” datasets.

**Extended Data Table 8:**
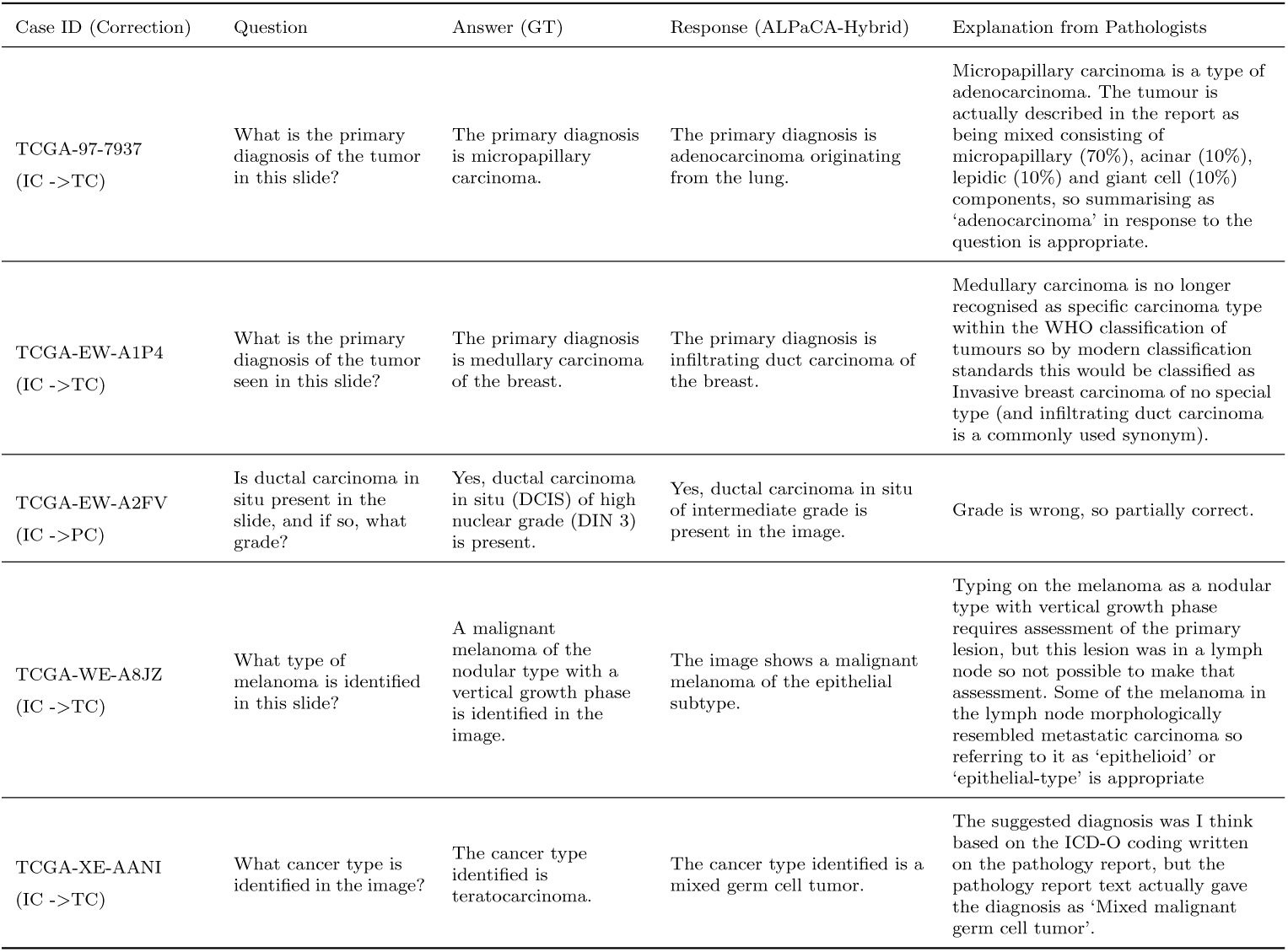
Examples of QA from the “General Slide-QA” dataset in the “Tumor Diagnosis and Classification” category, where expert pathologists corrected the judgments made by the GPT-4-based judger and provided detailed explanations.

**Extended Data Figure 1:**
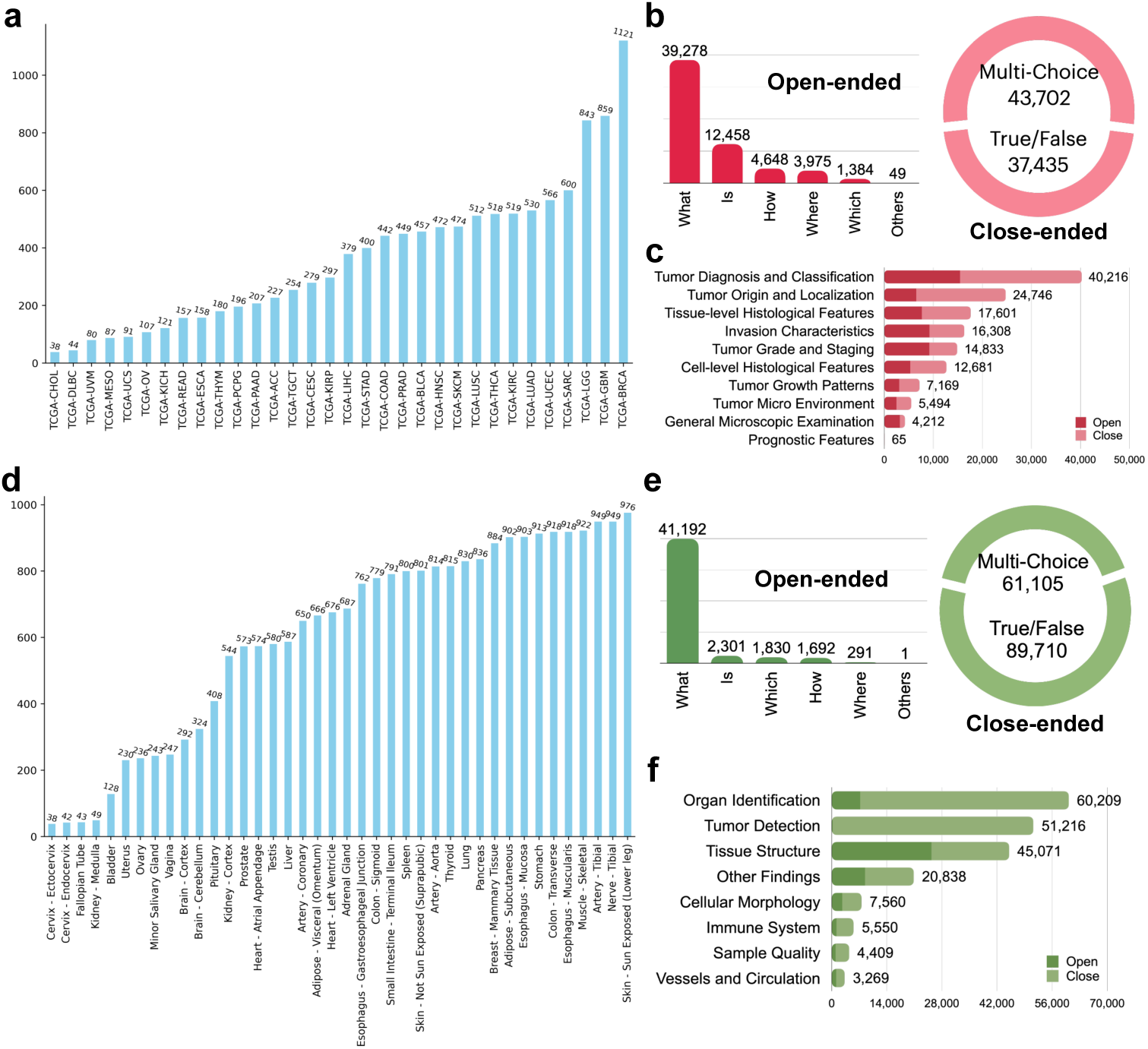
Dataset Distribution. **a**, Case distribution across different cancer projects in TCGA. **b**, Distribution of question types based on their nature in TCGA cases from the “General Slide-QA” dataset. **c**, Distribution of question categories in TCGA cases from the “General Slide-QA” dataset. **d**, Case distribution across different tissue sites in GTEx. **e**, Distribution of question types based on their nature in GTEx cases from the “General Slide-QA” dataset. **f**, Distribution of question categories in GTEx cases from the “General Slide-QA” dataset.

**Extended Data Figure 2:**
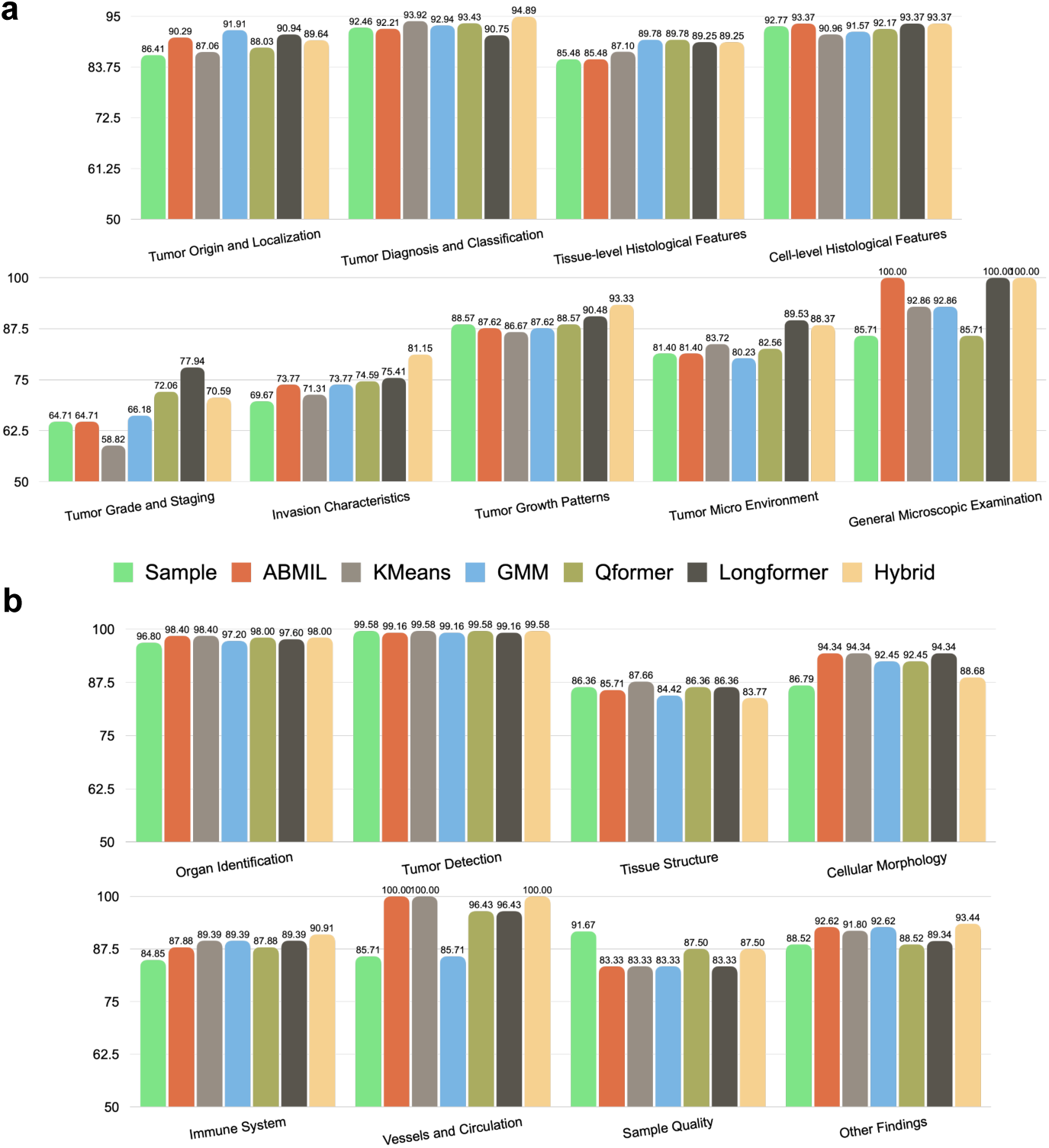
Close-ended performance (accuracy) of ALPaCA variants in the testset of “General Slide-QA”. **a**, Performance across various question categories in TCGA and GTEx. **b**, Performance across various question categories in GTEx cases.

**Extended Data Figure 3:**
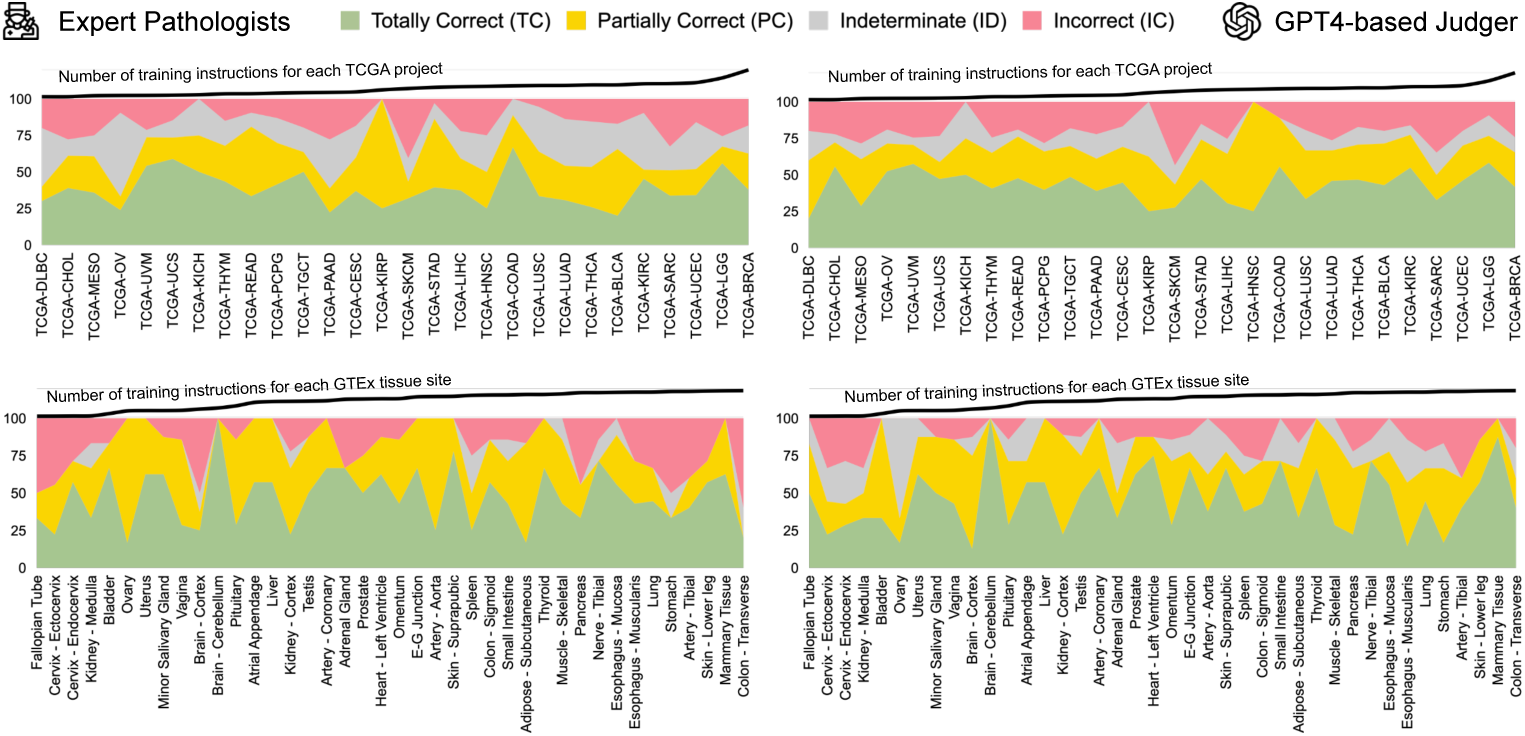
Open-ended performance (assessed by the GPT-4-based judger and expert pathologists) of ALPaCA-Hybrid across various cancer projects and tissue sites for TCGA and GTEx in the testset of “General Slide-QA”.

**Extended Data Figure 4:**
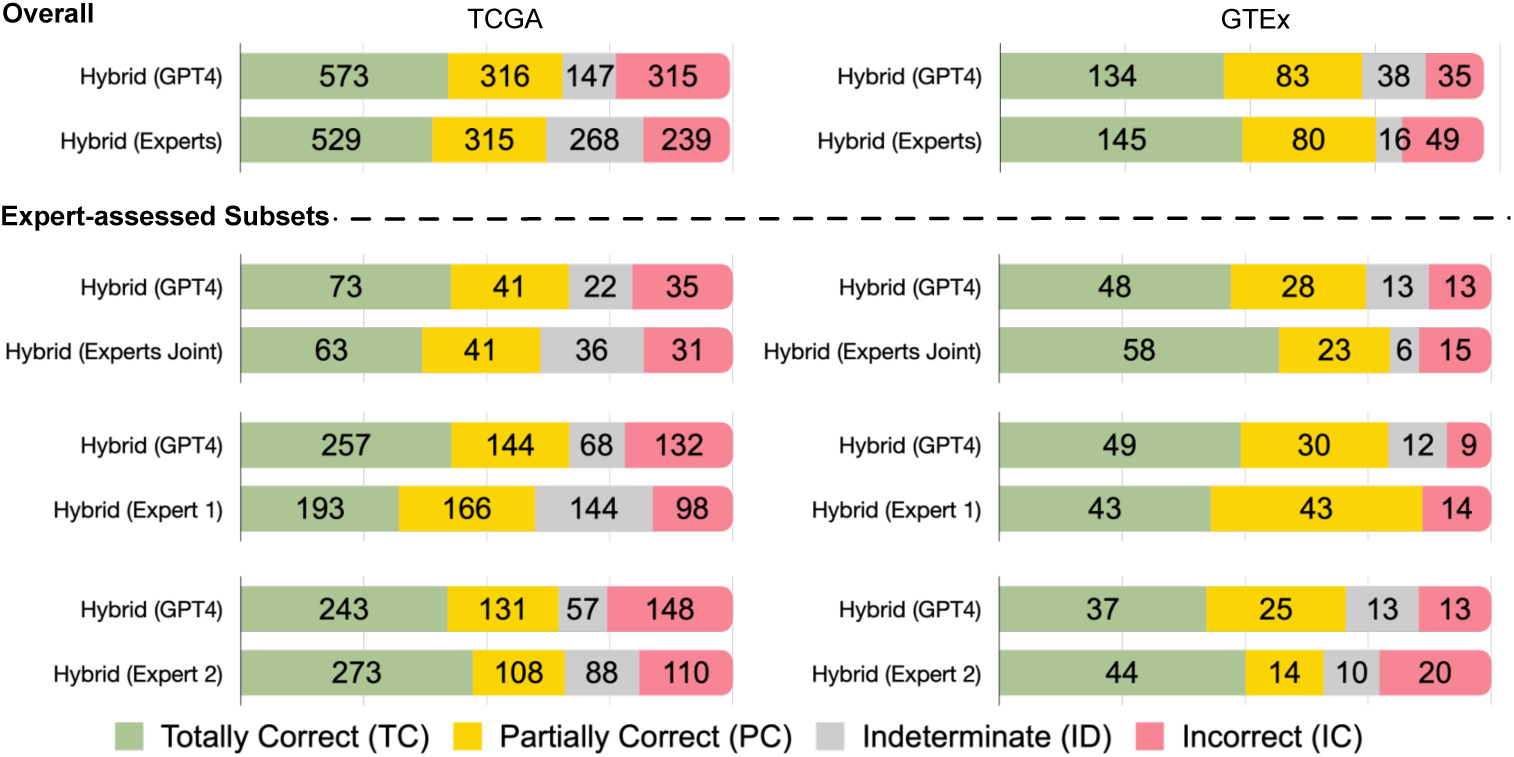
Open-ended response evaluation comparison of ALPaCA-Hybrid between the GPT-4-based judger and expert pathologists, further broken down by expert-assessed subsets for TCGA and GTEx in the testset of “General Slide-QA”.

**Extended Data Figure 5:**
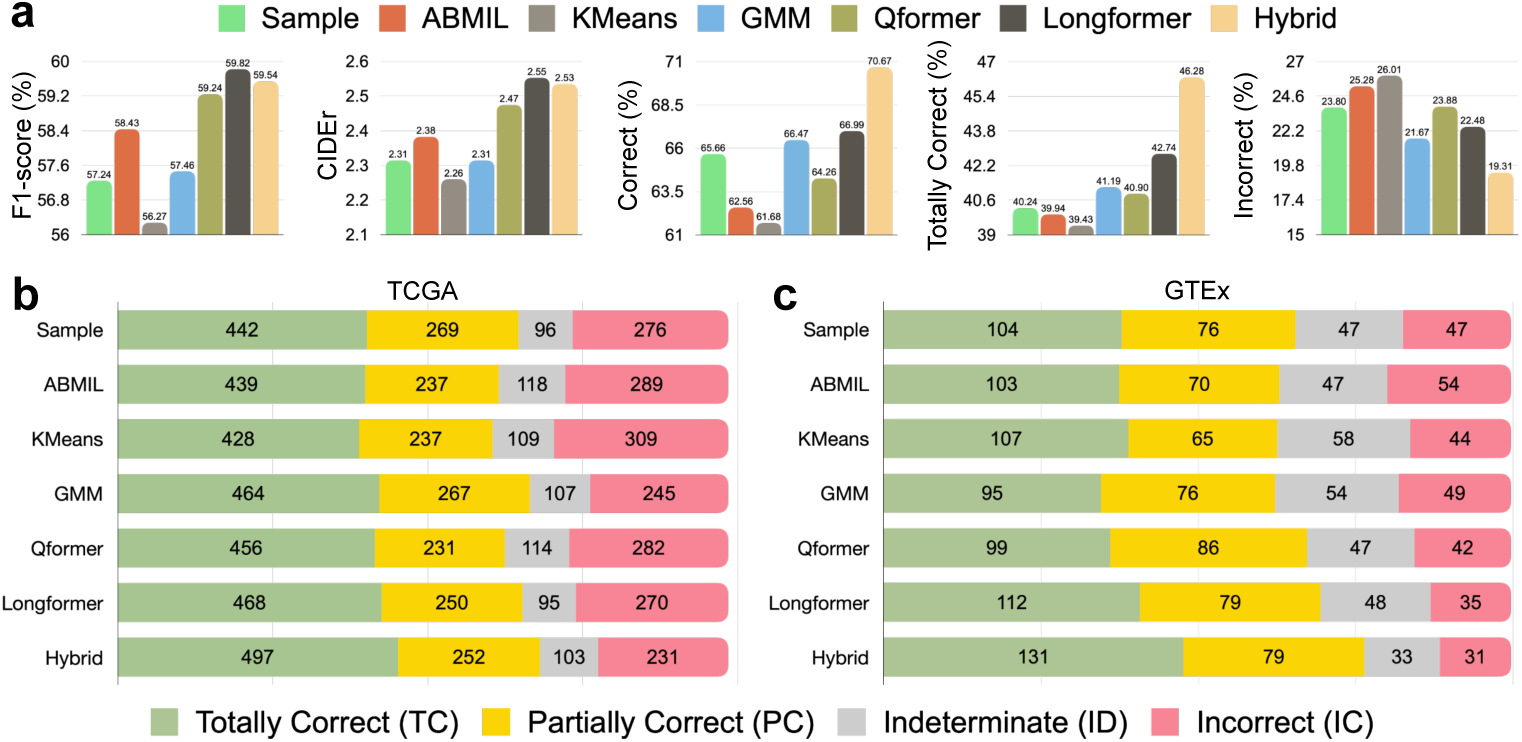
Open-ended performance of ALPaCA variants in the expert-curated testset of the “General Slide-QA”. **a**, the overall performance measured by word-level F1-score, CIDEr, and GPT-4-based judger. **b,c**, Detailed quantitative assessment results from the GPT-4-based judger for TCGA and GTEx.

