## Supplementary material for "ALPaCA: Adapting Llama for Pathology Context Analysis to enable slide-level question answering": prompt for gpt generation

### 1 Prompts

All prompts used for data curation and result evaluation are provided as follows:

- **TCGA Report Summary (GPT3.5 Turbo)**: *As a pathologist, please summarize the pathology report in one description paragraph. Content related to pathological examination must be retained as much as possible, and gross examination content is not required. If can not find any related content, return TBD.*
- **TCGA Report Refinement (GPT4o)**: *Imagine you are a pathologist looking at a primary tumor slide, you only know it is from which organ. To describe H&E stained WSI from the tumor center, please use the information from this pathology report. Note that the information here might involve multiple slides, you need to distinguish which of them can be observed from one single primary tumor slide only. Don't mention any information that cannot be obtained from the information of the primary tumor slide. Don't mention any information on metastasis, tumor measurements, tumor size, lymph node metastasis, tumor margins, and gross examination. The description should identify the original site, cancer type, specific subtype, stage (which system, like AJCC), and grade. Also include relevant descriptions from the primary tumor's H&E slide, if it exists. Do not describe anything that does not exist in the provided text, and summarize it into one paragraph.*
- **TCGA Open-ended Q&A Generation (GPT4o)**: *As a senior pathologist. You are given a text where a pathologist is describing a histopathology image. You are given the summary of a pathology slide. Generate well-defined question/answer pairs from these sentences. Consider the following requirements to generate questions and answers: - Only generate questions about information being seen in the image. - Do not generate questions from the context if the question is not explicitly asked. - Do not generate questions/answers based on information that can not be seen in the image being described like the patient's age, gender, medical history, or other studies/images outside of the current case. - If the*

*text does not explicitly provide the answer to a question, do not generate a question/answer pair. - Do not answer the questions from information outside of the given text. - Do not use phrases like "mentioned", "suggests" or "text" in the answers. Instead, refer to the information as being seen "in the image". - Do not reveal answers to the questions. In the returned JSON file, use key input to represent the question use the key output to represent the answer, and use qas as the key of a series of QA pairs.*

- **TCGA Close-ended (multi-choice) Q&A Generation (GPT4o)**: *You are a senior pathologist. You are given a text where a pathologist is describing a histopathology image. You are given the summary of a pathology slide. Generate concise multiple choice question answer pairs from the provided pathology slide summary. If there is enough information, you should at least ask the location or site of this image. When generating question-answer pairs, consider the following requirements: - Do not answer the questions as if you are reading a report but as if you are looking directly at the pathology slide. - Do not generate questions that need to be answered with numbers like scores or calculations. - Do not include right or left direction in any question and answer since you can not tell direction from the local image. - The key question should include the question. The key choices should include four choices. Each choice should be apparently diverse and contain a different format or pattern. Two choices should be apparently wrong with different formats. One choice should be similar to the correct answer. The key answer should include the correct choice. Provide choices with the numbers A, B, C, and D. Provide an answer with the proper number and the answer content. - In the generated choices, two of the choices should be apparently wrong with apparently different patterns. One choice should have a similar pattern with the correct answer serving as the confusing choice. - Do not generate questions/answers based on information that can not be seen in the image being described like the patients' age, gender, medical history, or other studies/images outside of the current case. - Do not answer the questions using information outside of the given text. - Do not use phrases like "mentioned", "suggests" or "text" in the answers. Instead, refer* *to the information as being seen "in the image". In the returned JSON file, use the key question to represent the question and four choices. Use the key answer to represent the corresponding answer and use qas to include a series of QA pairs. Don't forget to include two apparently irrelevant choices. Here is an example format 'qas':['question':'What is the colour of the leaf?' 'choices': 'A. Green. B. Red. C. Yellow. D. Bule','answer':'A. Green.' ].*

- **TCGA Close-ended (true or false) Q&A Generation (GPT4o)**: *You are a senior pathologist. You are given a text where a pathologist is describing a histopathology image. You are given the summary of a pathology slide. Generate yes-no question-answer pairs from the provided pathology slide summary. Consider the following requirements to generate questions and answers: - Do not answer the questions as if you are reading a report but like you are looking directly at the pathology slide. - Do not generate questions that are related to numbers like scores or calculations. - Change affirmation or denial tone to make sure more questions should be answered with 'No'. - The key question should include the question. The key answer should be only yes or no. Any other keys are not needed - Do not generate questions/answers based on information that can not be seen in the image being described like the patients' age, gender, medical history, or other studies/images outside of the current case. - Do not answer the questions using information outside of the given text. - Do not use phrases like "mentioned", "suggests" or "text" in the answers. Instead, refer to the information as being seen "in the image". - Do not reveal answers to the questions. In the returned JSON file, use the key question to represent the question. Use the key answer to represent the corresponding answer and use qas to include a series of QA pairs. Here is an example format 'qas':['question':'Is the colour of the leaf green?' 'answer':'Yes'] The summary content is as follows: content*
- **GTEx Notes Rephrasing (GPT4o)**: *Generate a one-sentence short descriptive report caption summarizing the key findings of a pathology assessment on the specific tissue, incorporating the major diagnostic categories related to the tissue condition, and providing a brief summary of the notable pathology information. Do not come to hypothetical*

*conclusions. Avoid providing content that can not be found in the original report. The provided report is from a non-tumor pathology slide. Do not include information about 'pieces', 'length', 'area', 'thickness', and other quantitive information. Generate the caption in a descriptive report tone like this slide is something; this slide reveals something. Rephrase the returned answer to remove the hyphen in the tissue column. In the returned JSON file, only one key called the 'caption' value should be the generated caption.*

- **GTEx Open-ended Q&A Generation (GPT4o)**: *As a senior pathologist, you are tasked with generating question/answer pairs based on the descriptions provided in a histopathology image summary. Please adhere to the following guidelines when formulating the questions and answers: 1. Limit the generation to two or three question/answer pairs per image caption, focusing on producing the most reliable pairs first. 2. Ensure that all captions and resultant question/answer pairs pertain exclusively to non-tumor pathology slides. 3. Questions should only pertain to specific, observable details in the image; avoid vagueness such as using terms like "notable." 4. Base questions solely on the content explicitly described in the image. Do not infer or assume additional information not presented. 5. Avoid creating question/answer pairs based on unobservable data such as the patient's demographic details, medical history, or external sources. 6. Formulate questions only if the text provides a clear answer; refrain from hypothesizing or assuming details not contained within the provided text. 7. In your answers, specify that details are "seen in the image," and avoid referencing the "text" or suggesting implications of the findings. 8. Craft questions in a way that they do not give away the answer. By following these guidelines, you will ensure that the question/answer pairs are accurate, specific, and strictly relevant to the given pathology slide images.*
- **GTEx Close-ended (multi-choice) Q&A Generation (GPT4o)**: *You are an experienced pathologist who is good at generating multiple-choice question-answer pairs. The provided report is from a non-tumor pathology slide. You should at least ask about the location or site of this image and whether this slide is non-tumor or contains a tumor. When*

*generating question-answer pairs, consider the following requirements: - Do not answer the questions as if you are reading a report but as if you are looking directly at the pathology slide. - Do not generate questions that need to be answered with numbers like scores or calculations. - Do not include right or left direction in any question and answer since you can not tell direction from the local image. - The key question should include the question. The key choices should include four choices. Each choice should be apparently diverse and contain a different format or pattern. Two choices should be apparently wrong with different formats. One choice should be similar to the correct answer. The key answer should include the correct choice. Provide choices with the numbers A, B, C, and D. Provide an answer with the proper number and the answer content. - In the generated choices, two of the choices should be apparently wrong with apparently different patterns. One choice should have a similar pattern with the correct answer serving as the confusing choice. - Do not generate questions/answers based on information that can not be seen in the image being described like the patients' age, gender, medical history, or other studies/images outside of the current case. - Do not answer the questions using information outside of the given text. - Do not use phrases like "mentioned", "suggests" or "text" in the answers. Instead, refer to the information as being seen in the image. In the returned JSON file, use the key question to represent the question and four choices. Use the key answer to represent the corresponding answer and use qas to include a series of QA pairs. Don't forget to include two apparently irrelevant choices. Here is an example format 'qas':['question':'What is the colour of the leaf?' 'choices': 'A. Green. B. Red. C. Yellow. D. Bule', 'answer': 'A. Green.'].*

- **GTEx Close-ended (true or false) Q&A Generation (GPT4o)**: *You are an experienced pathologist who is good at generating multiple-choice question-answer pairs. The provided report is from a non-tumor pathology slide. You should at least ask the location or site of this image and whether this slide is non-tumor or contains tumor.' When generating question-answer pairs, consider the following requirements: - Do not generate questions related to "pieces" in the clinical notes. - Generate the most clear and certain* *question pair. Do not generate questions that are similar to each other. - Do not answer the questions as if you are reading a report but as if you are looking directly at the pathology slide. - Do not generate questions that need to be answered with numbers like scores or calculation or counting results. - Do not include right or left direction in any question or answer since you can not tell direction from the local image. - Change affirmation or denial tone to make sure answer "Yes" and "No" are evenly distributed. - The key question should include the question. The key answer should be only yes or no. No other keys are needed - Do not generate questions/answers based on information that can not be seen in the image being described like the patients' age, gender, medical history, or other studies/images outside of the current case. - Do not answer the questions using information outside of the given text. - Do not use phrases like "mentioned", "suggests" or "text" in the answers. Instead, refer to the information as being seen "in the image". In the returned JSON file, use the key question to represent the question. Use the key answer to represent the corresponding answer and use qas to include a series of QA pairs. Here is an example format 'qas':['question':'Is the color of the leaf green?' 'answer':'Yes'] The summary content is as follows: report*

- **TCGA-BRCA Open-ended Q&A Generation (GPT4o)**: *You will be given a Dict containing detailed information about breast cancer. Generate well-defined question/answer pairs from these key-value pairs. Try using only the original content and avoid adding hallucination. You can only make a little rephrase to avoid repetition. In the returned JSON file, use key input to represent the question and use key output to represent the answer and use qas as the key of a series of QA pairs. The summary content is as follows: content*
- **TCGA-BRCA Close-ended (multi-choice) Q&A Generation (GPT4o)**: *You will be given a Dict containing detailed information about breast cancer. Generate well-defined question/answer pairs from these key-value pairs. Try using only the original*

*content and avoid adding hallucination. You can only make a little rephrase to avoid repetition. When generating question-answer pairs, consider the following requirements: - Do not answer the questions as if you are reading a report but as if you are looking directly at the pathology slide. - Do not include right or left direction in any question and answer since you can not tell direction from the local image. - Do not generate questions and answers that should be answered with numbers like how many counting results. - The key question should include the question. The key choices should consist of four choices. Each choice should be apparently diverse and contain a different format or pattern. Two choices should be apparently wrong with different formats. One choice should be similar to the correct answer. The key answer should include the correct choice. Provide choices with the numbers A, B, C, and D. Provide an answer with the proper number and the answer content. - In the generated choices, two of the choices should be apparently wrong with apparently different patterns. One choice should have a similar pattern with the correct answer serving as the confusing choice. - Do not generate questions/answers based on information that can not be seen in the image being described like the patients' age, gender, medical history, or other studies/images outside of the current case. - Do not answer the questions using information outside of the given text. - Do not use phrases like "mentioned", "suggests" or "text" in the answers. Instead, refer to the information as being seen "in the image". In the returned JSON file, use the key question to represent the question and four choices. Use the key answer to represent the corresponding answer and use qas to include a series of QA pairs. Don't forget to include two apparently wrong choices. Here is an example format 'qas':['question':'What is the color of the leaf?' 'choices': 'A. Green. B. Red. C. Yellow. D. Bule','answer':'A. Green. '] The summary content is as follows: content*

- **TCGA-BRCA Close-ended (true or false) Q&A Generation (GPT4o)**: *You will be given a Dict containing detailed information about breast cancer. Generate well-defined question/answer pairs from these key-value pairs. When generating question-answer pairs, consider the following requirements: - Generate the most clear and certain question* *pair. Generate at most 5 certain question-answer pairs for each summary. Do not generate questions that are similar to each other. - Do not answer the questions as if you are reading a report but as if you are looking directly at the pathology slide. - Do not generate questions that need to be answered with numbers like scores or calculation or counting results. - Do not include right or left direction in any question or answer since you can not tell direction from the local image. - Change affirmation or denial tone to make sure answers "Yes" and "No" are evenly distributed. - The key question should include the question. The key answer should be only yes or no. No other keys are needed - Do not generate questions/answers based on information that can not be seen in the image being described like the patients' age, gender, medical history, or other studies/images outside of the current case. - Do not answer the questions using information outside of the given text. - Do not use phrases like "mentioned", "suggests" or "text" in the answers. Instead, refer to the information as being seen "in the image". In the returned JSON file, use the key question to represent the question. Use the key answer to represent the corresponding answer and use qas to include a series of QA pairs. Here is an example format 'qas':['question':'Is the color of the leaf green?' 'answer':'Yes'] The summary content is as follows: content*

- **Open-ended Q&A evaluation (GPT4o)**: *You will be provided with a question, a ground truth (GT) answer, and a model-generated result related to a pathology case. Your task is to evaluate the correctness of the model's predicted result. You are the final judge and must follow these instructions: 1. If the model's result and the GT answer convey similar or equivalent information, it is correct, even if the wording or structure differs. 2. If the model's result is broader but includes the GT answer or its key elements, it should still be considered correct. 3. If part of the model's result matches part of the key information from the GT answer, the result is partially correct. 4. If the model's result and the GT answer do not overlap, but the result could plausibly serve as an alternate answer to the question, it is uncertain. 5. Since the GT answer is derived from a pathology report that summarizes information from multiple slides, it may not always be accurate or fully representative of* *the specific slide in question. Only GT answers directly related to a definitive diagnosis can be fully relied upon. 6. For morphology-related questions, the model's result should only be considered incorrect if it directly and completely contradicts the GT answer, such that both cannot plausibly coexist on the same slide. Provide your evaluation as a single number: 2 if the model's result is correct, 1 if it is partially correct, 0 if it is uncertain, -1 if it is incorrect. Ensure your assessment reflects the clinical context and the accuracy of the provided content.*
